# Allelic effects on *KLHL17* expression likely mediated by JunB/D underlie a PDAC GWAS signal at chr1p36.33

**DOI:** 10.1101/2024.09.16.24313748

**Authors:** Katelyn E. Connelly, Katherine Hullin, Ehssan Abdolalizadeh, Jun Zhong, Daina Eiser, Aidan O’Brien, Irene Collins, Sudipto Das, Gerard Duncan, Pancreatic Cancer Cohort Consortium, Pancreatic Cancer Case-Control Consortium, Stephen J. Chanock, Rachael Z. Stolzenberg-Solomon, Alison P. Klein, Brian M. Wolpin, Jason W. Hoskins, Thorkell Andresson, Jill P. Smith, Laufey T. Amundadottir

## Abstract

Pancreatic Ductal Adenocarcinoma (PDAC) is the third leading cause of cancer-related deaths in the U.S. Both rare and common germline variants contribute to PDAC risk. Here, we fine-map and functionally characterize a common PDAC risk signal at 1p36.33 (tagged by rs13303010) identified through a genome wide association study (GWAS). One of the fine-mapped SNPs, rs13303160 (r^2^=0.93 in 1000G EUR samples, OR=1.23, *P* value=2.74×10^−9^) demonstrated allele-preferential gene regulatory activity *in vitro* and allele-preferential binding of JunB and JunD *in vitro* and *in vivo*. Expression Quantitative Trait Locus (eQTL) analysis identified *KLHL17* as a likely target gene underlying the signal. Proteomic analysis identified KLHL17 as a member of the Cullin-E3 ubiquitin ligase complex in PDAC-derived cells. *In silico* differential gene expression analysis of the GTExv8 pancreas data suggested an association between lower KLHL17 (risk associated) and pro-inflammatory pathways. We hypothesize that KLHL17 may mitigate inflammation by recruiting pro-inflammatory proteins for ubiquitination and degradation thereby influencing PDAC risk.

## Introduction

Pancreatic cancer is currently the third leading cause of cancer-related deaths and is expected to move to second place by 2030 in the United States.^1^ Pancreatic ductal adenocarcinoma (PDAC) comprises over 90% of pancreatic cancer cases. While its survival rate has improved over the years, detection, prevention, and treatment of PDAC remains a challenge.^2^ Epidemiological factors known to increase risk to PDAC include Type 2 diabetes, pancreatitis, and obesity.^3^ Additionally, both rare high-risk and common, low effect size germline variants are known to contribute to PDAC susceptibility.^4–8^

The Pancreatic Cancer Cohort Consortium and Pancreatic Cancer Case-Control Consortium have sought to identify common germline variants that influence risk of PDAC through genome-wide association studies (GWAS). Previous GWAS phases, PanScan I,^5^ II,^6^ and III^7,8^ and PanC4,^9^ have identified 17 independent risk signals for PDAC. A 2018 meta-analysis of these four studies (9,040 cases and 12,496 controls) and TaqMan replication using samples from the PANcreatic Disease ReseArch (PANDoRA) consortium (2,737 cases and 4752 controls) uncovered five new signals.^4^ One of the newly identified loci was at chr1p36.33. The initial meta-analysis of PanScan I, II, III and PanC4 identified 1p36.33 (tagged by single nucleotide polymorphism (SNP) rs13303010; *P* = 7.3×10^−7^; odds ratio (OR) = 1.20; 95% confidence interval (CI) = 1.12-1.29) as a suggestive risk locus for PDAC (Table 1). A meta-analysis including samples from the PANDoRA consortium (11,537 cases and 17,107 controls) improved the signal beyond the GWAS significance threshold (rs13303010, OR = 1.26, 95% CI 1.19-1.35, *P* value = 8.36×10^−14^) (Table 1),^4^ further supporting 1p36.33 as a PDAC risk locus.

**Table 1:** Summary statistics for rs13303010 on 1p36.33 PDAC risk locus.

| <sup>a</sup>Marker, <sup>b</sup>alleles, <sup>c</sup>chr, <sup>d</sup>location, and <sup>e</sup>gene | Study | MAF |  | Subjects |  | *P*-value | Allelic OR (95% CI) |
| --- | --- | --- | --- | --- | --- | --- | --- |
|  |  | Controls | Cases | Controls | Cases |  |  |
| rs13303010 (G, A) | 2018 Meta Analysis<sup>4</sup> |  |  | 12,496 | 9,040 | 7.3x10<sup>-7</sup> | 1.20 (1.12–1.29) |
| 1p36.33 (894,573) | PANDoRA<sup>4</sup> | 0.10 | 0.14 | 4,611 | 2,497 | 6.0x10<sup>-10</sup> | 1.45 (1.33–1.57) |
| *NOC2L* | Meta Analysis + PANDoRA<sup>4</sup> |  |  | 17,107 | 11,537 | 8.36x10<sup>-14</sup> | 1.26 (1.19–1.35) |
|  | UK Biobank | 0.09 | 0.12 | 9,399 | 1,066 | 1.05x10<sup>-5</sup> | 1.42 (1.21-1.65) |
|  | Meta Analysis<sup>4</sup> + UK Biobank |  |  | 21,895 | 10,106 | 2.09x10<sup>-10</sup> | 1.24 (1.16-1.32) |
| Summary statistics from previously published PDAC GWAS and for the new meta-analysis presented here including UK Biobank PDAC cases and controls for rs13303010 including allele frequencies in cases and controls, *P*-value, and odds ratio (OR) with the 95% confidence intervals (CI). <sup>a</sup> rsID; SNP ID number, <sup>b</sup>Minor, major alleles; <sup>c</sup>location; <sup>d</sup>Position (Genome build hg19); <sup>e</sup>Nearest gene |  |  |  |  |  |  |  |

Most GWAS signals map to non-coding, regulatory regions of the genome and are hypothesized to influence disease risk through allele specific changes in gene expression.^10^ 1p36.33 maps to a gene dense region, with the tag SNP rs13303010 lying within the first intron of *NOC2L*. Here, we applied fine-mapping methods to identify candidate functional variants for *in vitro* testing. We subsequently identified rs13303160 as a functional variant likely mediating the expression of *KLHL17* through allele-preferential binding of JunB and JunD transcription factors. Work to characterize *KLHL17’s* function in PDAC risk suggested a role for KLHL17 in mitigating inflammation through protein homeostasis.

## Results

### Fine-mapping of the chr1p36.33 PDAC risk locus

To identify candidate functional variants at the 1p36.33 risk locus, we first performed a meta-analysis using GWAS data from PanScan I-III, PanC4 as well as an additional 1,066 PDAC cases and 9,399 control subjects from the UK Biobank (UKBB).^11^ This resulted in a total of 10,106 cases and 21,895 control subjects with imputed GWAS data. In this analysis, rs13303010 remained the most significant SNP at 1p36.33 (OR=1.24, 95% CI 1.16-1.32, *P*=2.09×10^−10^) (Figure 1, Table 1).

**Figure 1:**
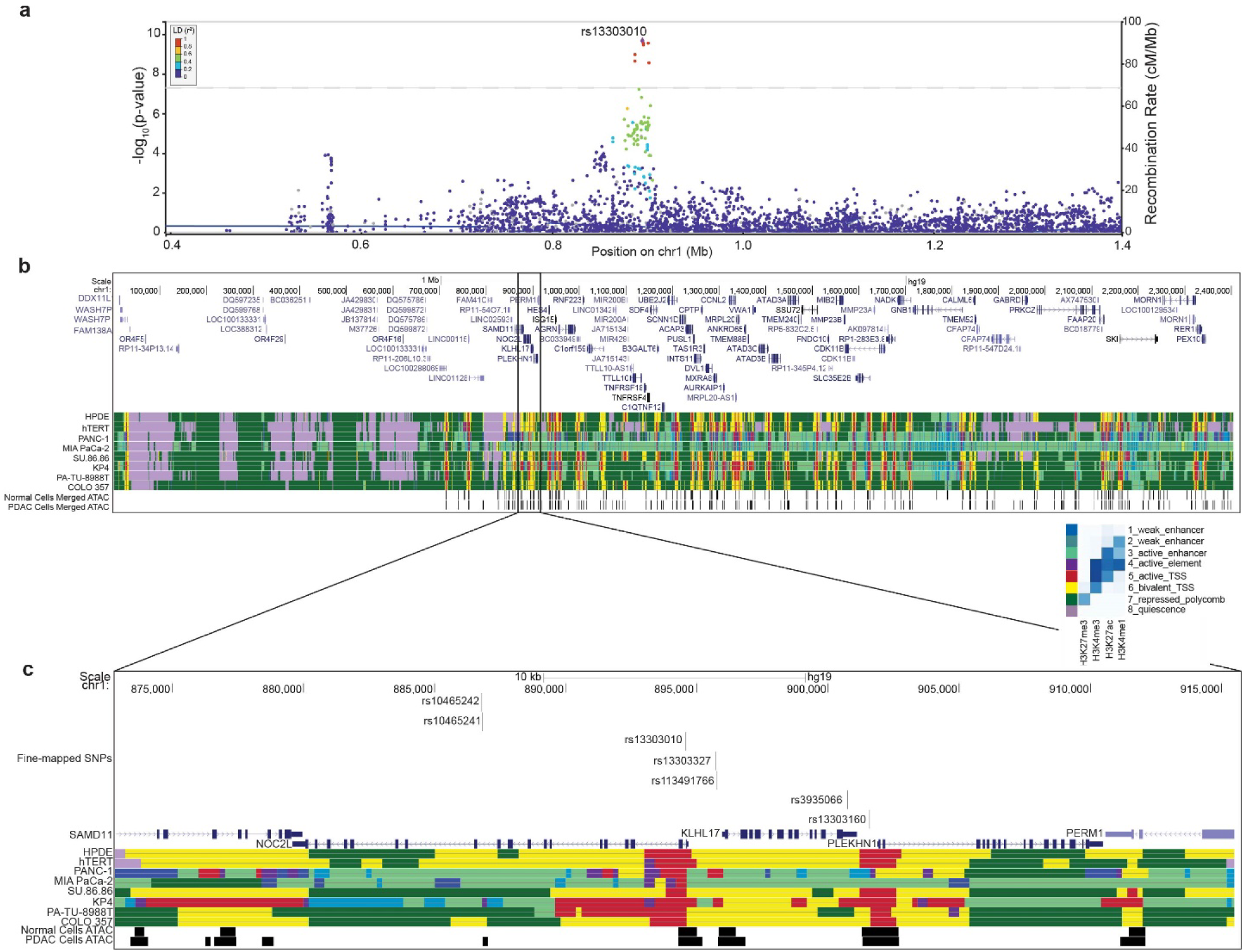
Overview of chr1p36.33 PDAC risk locus. a) Locus Zoom plot of the variants identified in the meta-analysis, colors are indicative of the LD r2 in reference to the lead SNP b) UCSC genome browser view of 1p36.33 with ChromHMM and ATAC-seq annotations in PDAC and normal-derived duct epithelial cell lines c) Zoomed in UCSC browser showing the candidate functional variants and nearby genes

To identify credible causal variants (CCVs) underlying this association signal, we applied fine-mapping approaches to the summary statistics for our latest meta-analysis. We implemented the Bayesian approach Sum of Single Effects Linear Regression (SuSiE)^12^ to identify credible sets of CCV with 90% confidence. SuSiE identified one credible set with five variants (Table 2, highlighted). Additionally, we applied a likelihood ratio (LLR < 1:100) and linkage disequilibrium (LD r^2^ > 0.8) threshold. This identified two additional CCVs (Table 2). To be inclusive, we moved all seven variants forward for *in vitro* functional analysis.

**Table 2:** Summary statistics and fine-mapping for the top 10 SNPs in UK Biobank meta-analysis.

| <sup>a</sup>Marker, <sup>b</sup>alleles, <sup>c</sup>location | OR (95% CI) | *P*-value | LD *r*<sup>2</sup> | LLR | SuSiE PIP | Chromatin Accessibility |
| --- | --- | --- | --- | --- | --- | --- |
| rs13303010 (G, A) 1:894,573 | 1.24 (1.16-1.32) | 2.09x10<sup>-10</sup> | -- | 1.00 | 0.27 | Yes |
| rs3935066 (G, A) 1:900,730 | 1.25 (1.17-1.35) | 2.74x10<sup>-10</sup> | 0.89 | 1.30 | 0.22 | No (>500bp) |
| rs113491766 (A, AG)<br>1:895,755 | 0.81 (0.76-0.86) | 2.85x10<sup>-10</sup> | 1 | 1.35 | 0.20 | ~56 bp away |
| rs13303327 (G, A) 1:895,706 | 1.24 (1.16-1.32) | 3.38x10<sup>-10</sup> | 1 | 1.60 | 0.18 | ~110bp away |
| rs10465241 (C, T) 1:886,817 | 1.24 (1.16-1.32) | 1.03x10<sup>-9</sup> | 0.90 | 4.74 | 0.06 | ~22 bp |
| rs10465242 (G, A) 1:886,788 | 1.23 (1.15-1.32) | 2.23x10<sup>-9</sup> | 0.90 | 10.07 | 0.03 | ~52 bp |
| rs13303160 (G, A) 1:901,559 | 1.23 (1.15-1.32) | 2.74x10<sup>-9</sup> | 0.93 | 12.31 | 0.02 | Yes |
| rs133032957 (G, A) 1:891,021 | 1.26 (1.16-1.37) | 5.92x10<sup>-8</sup> | 0.52 | 243.62 | 1.34x10<sup>-3</sup> | -- |
| rs4970445 (G, A) 1:891,021 | 1.23 (1.14-1.33) | 1.53x10<sup>-7</sup> | 0.58 | 610.81 | 5.37x10<sup>-4</sup> | -- |
| rs7524174 (G, A) 1:902997 | 1.19 (1.11-1.27) | 3.94x10<sup>-7</sup> | 0.57 | 1523.22 | 2.32x10<sup>-4</sup> | -- |

Summary statistics of the top 10 for the newest meta-analysis including PDAC UK Biobank cases and controls including OR and 95% CI, *P* value, linkage disequilibrium (LD) *r*<sup>2</sup> reported relative to the tag SNP based on the European 1000Genomes reference, likelihood ratio (LLR), SuSiE posterior inclusion probability (PIP), and proximity to accessible chromatin. SNPs highlighted in yellow were identified in the SuSiE credible set of variants. All variants above the dotted line met the fine-mapping criteria for functional follow-up. Accessible chromatin is based on the ATAC-seq data generated in PDAC-derived or normal-derived pancreas epithelial cell lines. <sup>a</sup>rsID; <sup>b</sup>Minor, major alleles; <sup>c</sup>chr:position (Genome build hg19)

Most GWAS variants are noncoding and thought to affect gene expression of target genes in an allele specific manner. As such, variants have been shown to be enriched in active gene regulatory elements indicated by posttranslational modifications of histones (H3K4me3, H3K4me1, H3K27ac) and accessible chromatin.^13–16^ We examined the set of statistically fine-mapped variants in the context of pancreas gene regulatory elements, using a ChromHMM 8-state model and ATAC-seq open chromatin data we generated in PDAC and normal-derived pancreas cell lines.^17,18^ All these variants lie within active and bivalent transcriptional start sites and active enhancer elements (Fig. 1B). Two variants lie within regions of open chromatin (Fig. 1C, Table 2) further supporting the fine-mapped variants as candidate functional SNPs influencing gene expression in *cis* or *trans*.

### Assessing allele-preferential binding and gene regulatory activity of candidate functional variants

To identify functional variants underlying this GWAS signal, we first sought to identify variants that exhibited allele-preferential transcription factor (TF) binding. We tested the set of seven fine-mapped variants at chr1p36.33 in electrophoretic mobility shift assays (EMSAs) using nuclear extracts from the PANC-1 pancreatic cancer cell line. Three of the seven variants demonstrated allele-preferential protein binding: rs13303010, rs13303327, and rs13303160 (Fig. 2). The GWAS tag SNP, rs13303010 showed preferential binding to the protective alternate allele (A) (Fig. 2a). Additionally, we observed consistent allele-preferential binding with the protective alternate allele (A) at rs13303327 (Fig. 2b). The third variant, rs13303160, exhibited preferential binding to the risk-increasing reference (G) allele (Fig. 2c). These preferential binding patterns for all SNPs were also observed with MIA PaCa-2 (Supplementary Fig. 1a-c) and HeLa nuclear lysate (Supplementary Fig. 1d, e).

**Figure 2:**
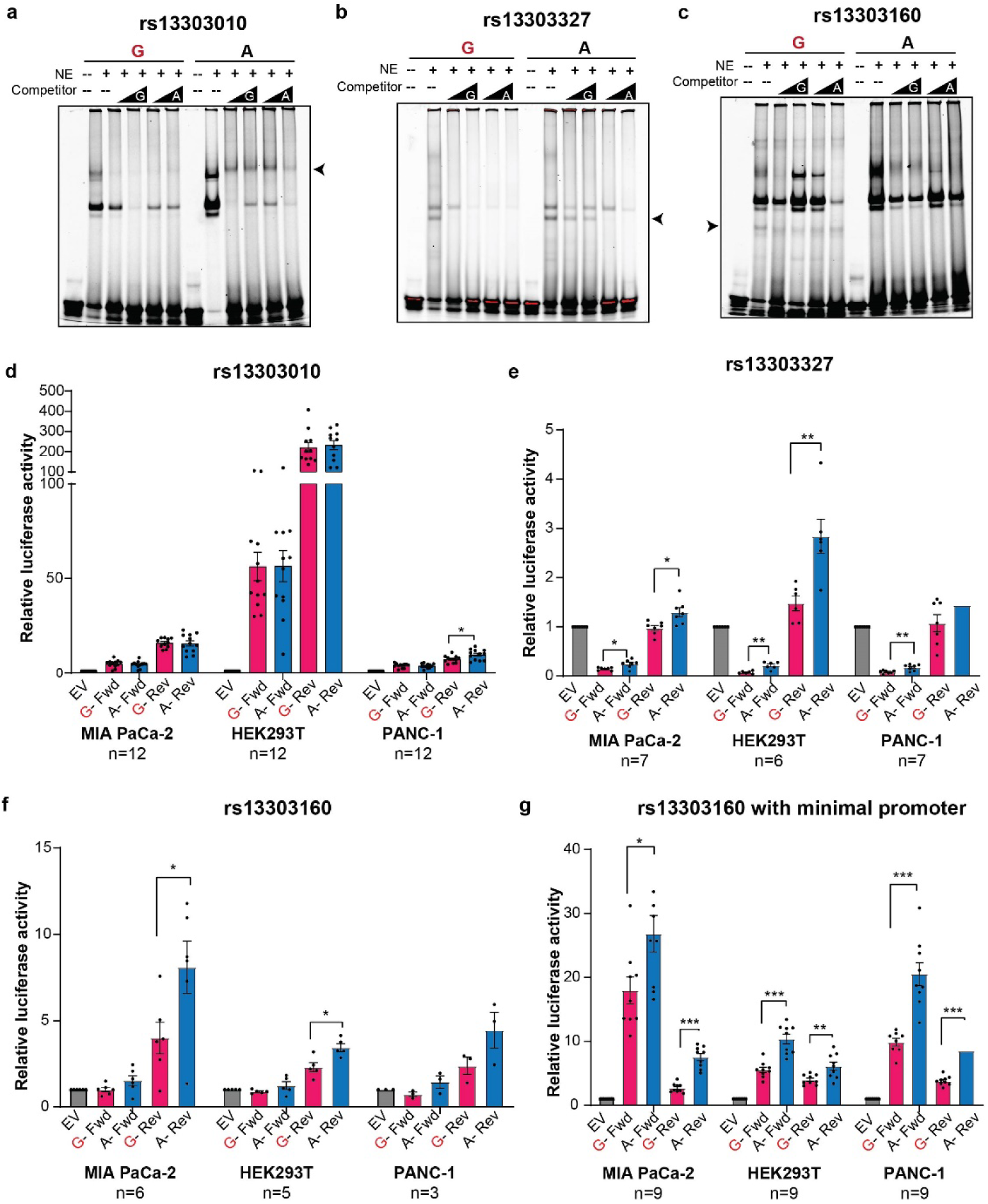
Identification of allele-preferential binding and activity using EMSA and luciferase reporter assays. a-c) Representative EMSA with PANC-1 nuclear extract and fluorescently labelled 31bp oligonucleotides with the variant, rs13303010, rs13303327, rs13303160, respectively centered in the middle. Competitor is the same sequence with no fluorescent label in excess (50, 100X); d-f) Luciferase reporter assay using rs13303010, rs13303327, and rs13330160, respectively and the surrounding sequence as a promoter to the luciferase gene in three cell lines, number of biological replicates are indicated below the cell line name; g) Luciferase assay for rs13303160 and surrounding sequence as an enhancer upstream of a minimal promoter and luciferase gene in three cell lines. The number of replicates is indicated below the cell line name. The risk allele is colored in red. For luciferase, the forward and reverse orientation of the sequence was used. Error bars represent the standard error of the mean (SEM) and significance was determined by an unpaired, two-tailed t-test; * *P* <0.05; ** *P* < 0.01, *** *P* < 0.001.

We next took the three variants with allele-preferential binding forward to evaluate allele-preferential gene regulatory activity using luciferase reporter activity assay. As these variants lie near active transcriptional start sites (TSS) (Fig. 1c), we first tested allele-preferential promoter activity by placing 141-201 base pair (bp) sequences centered on the variant of interest (see Methods) into a luciferase vector without a minimal promoter (in the pGL4.14 vector). We observed strong promoter activity for the rs13303010 constructs compared to the empty vector (EV) but minimal allele-preferential luciferase activity in MIA PaCa-2, PANC-1 and HEK293T cell (Fig. 2d). The second SNP, rs13303327 demonstrated allele-preferential luciferase activity with the alternate (A) allele having a stronger regulatory effect (Fig. 2e). The third SNP, rs13303160, exhibited strong promoter activity with the alternate (A) allele demonstrating a specific allele-preferential effect in the reverse orientation in the MIA PaCa-2 and HEK293T cells (Fig. 2f).

While rs13303010 and rs13303327 lie in a 1,328 bp region between the transcriptional start sites (TSS) of the *NOC2L* and *KLHL17* genes, rs13303160 is located 360 bps downstream of the 3’UTR of *KLHL17* and 303 bps upstream of the TSS for *PLEKHN1* (Fig. 1c), suggesting it could influence promoter and/or enhancer activity at this locus. We therefore tested the sequence surrounding rs13303160 as an enhancer upstream of a minimal promoter and the luciferase gene (in the pGL4.23 vector). As an enhancer element, the rs13303160 constructs demonstrated strong enhancer activity in all three cell lines with the alternate (A) allele exhibiting stronger activity (Fold change (FC) 1.5-2.8, *P*= 1×10^−6^-0.02; Fig. 2g). Thus, through EMSA and luciferase assays, we narrowed the set of seven fine-mapped candidate functional variants down to two (rs13303327 and rs13303160) that each demonstrate allele-preferential protein binding and gene regulatory activity.

### Identifying allele-preferential protein binding for rs13303327 and rs13303160

To identify TFs potentially mediating the allele-preferential regulatory activity we observed, we performed an *in silico* motif analysis using PERFECTOS-APE.^19^ This analysis predicts TF binding potential for both alleles of a variant and provides a *P*-value for the estimated strength of binding. We then calculated the fold-change between *P*-values as a proxy for the binding affinity change between alleles.

For rs13303327, we identified several E74-like factors (ELF), members of the E-twenty-six (ETS) family of transcription factors, having a 13-24-fold difference in binding *P*-values between the A and G alleles (Table 3, Supplementary Table 1). The ELF transcription factors recognize the motif GGAA (Figure 3a) which is disrupted by the risk allele-G at rs13303327 by replacing the last A with a G.

**Figure 3:**
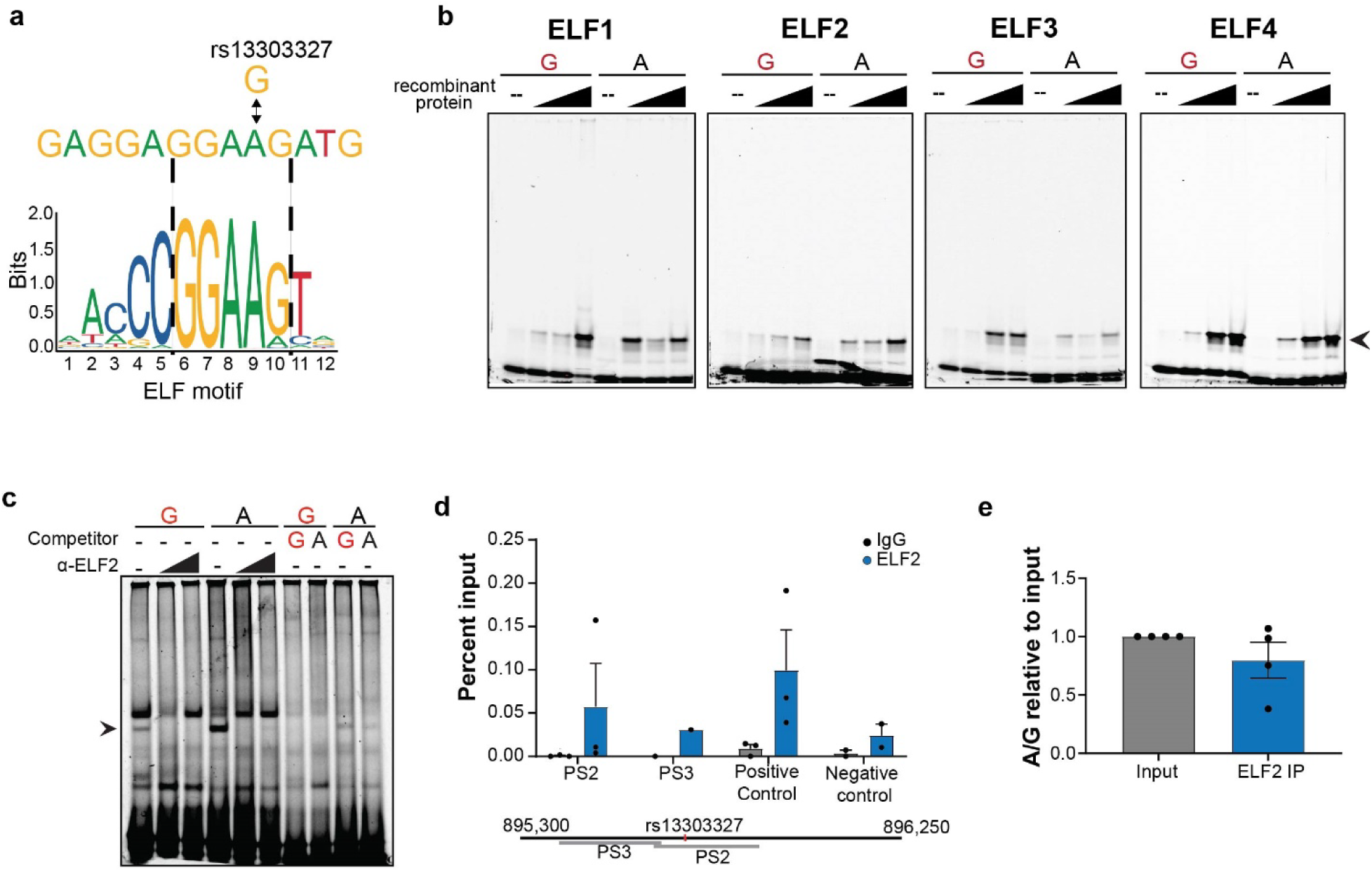
Allele-preferential binding of ELF to rs13303327 *in vitro* and *in vivo*. a) *in silico* TF binding motif prediction for rs13303327; b) EMSA with increasing amounts of recombinant ELF proteins and fluorescently labeled oligonucleotide; c) EMSA supershift with an antibody against ELF2 using PANC-1 nuclear lysate; d) ChIP-qPCR for ELF2 in Hs766T PDAC cell line with primers near or encompassing the SNP. The positive control is a documented region from an ELF2 ChIP-seq in K562 and negative control is from a quiescent region of 1p36.33. Percent input enrichment was quantitatively determined using a standard curve derived from input DNA as described in the ActiveMotif protocol); e) TaqMan genotyping of enriched ChIP-qPCR DNA; A to G ratio was calculated in relative to the input ratio. For ChIP-qPCR, all IPs were performed in quadruplicate, but IPs that did not enrich are not shown. Error bars represent SEM. Unpaired, two-tailed t-tests were performed; nothing was deemed significant (*P* <0.05)

**Table 3:** Predicted TFs with allelic binding preferences for rs13303327 and rs13303160.

| rsID | Motif | Allele 1 | Allele 2 | *P*-value Allele 1 | *P*-value Allele 2 | *P*-value Fold Change |
| --- | --- | --- | --- | --- | --- | --- |
| rs13303327 | ELF1 | G | A | 2.27x10<sup>-3</sup> | 1.28x10<sup>-4</sup> | 17.74 |
| rs13303327 | ELF2 | G | A | 2.08x10<sup>-3</sup> | 8.70x10<sup>-5</sup> | 23.91 |
| rs13303327 | ELF3 | G | A | 1.81x10<sup>-3</sup> | 1.31x10<sup>-4</sup> | 13.83 |
| rs13303327 | ELF5 | G | A | 6.56x10<sup>-4</sup> | 2.66x10<sup>-5</sup> | 24.64 |
| rs13303160 | FOSB | G | A | 1.18x10<sup>-4</sup> | 2.93x10<sup>-5</sup> | 40.11 |
| rs13303160 | FOS | G | A | 5.73x10<sup>-4</sup> | 2.74x10<sup>-5</sup> | 20.86 |
| rs13303160 | FOSL1 | G | A | 4.32x10<sup>-4</sup> | 4.94x10<sup>-6</sup> | 87.31 |
| rs13303160 | FOSL2 | G | A | 8.16x10<sup>-4</sup> | 2.15x10<sup>-5</sup> | 37.93 |
| rs13303160 | JUNB | G | A | 8.00x10<sup>-4</sup> | 2.47x10<sup>-5</sup> | 32.36 |
| rs13303160 | JUND | G | A | 9.47x10<sup>-4</sup> | 2.28x10<sup>-5</sup> | 41.63 |
| rs13303160 | JUN | G | A | 8.40x10<sup>-4</sup> | 2.68x10<sup>-5</sup> | 31.37 |

To validate TF binding predictions for rs13303327, we performed EMSAs with recombinant ELF1, ELF2, ELF3, and ELF4 proteins to assess allele-preferential binding *in vitro* (Fig. 3b). ELF2 consistently demonstrated preferential binding to the A allele as compared to the G allele (Fig. 3b, Supplementary Fig. 2). ELF1, 3 and 4 did not demonstrate consistent or differences in allelic binding (Fig.3b). Additionally, including an ELF2 antibody led to a higher mobility band (supershift) further confirming ELF2 as the protein responsible for the allele-preferential band in the original EMSAs (Fig. 3c).

To assess if ELF2 is enriched at rs13303327 in an allele-preferential manner in the context of the native DNA, we performed chromatin immunoprecipitation followed by quantitative PCR (ChIP-qPCR) in two pancreatic cancer cell lines heterozygous at this SNP (Hs766T and SW1990). We were unable to enrich ELF2 at rs13303327 or a predicted positive control region from the SW1990 PDAC cell line. In Hs766T cells, we observed minimal enrichment of ELF2 relative to the IgG and negative control region (Fig. 3d).

We then assessed allele-preferential enrichment of the ChIP DNA using a TaqMan genotype probe for rs13303327 and observed an enrichment of ELF2 over IgG; however, an allele-preferential enrichment compared to the input DNA was not noted (Fig. 3e). Based on these results, we conclude that ELF2 does not bind rs13303327 in an allele-preferential manner in PDAC cell lines.

For rs13303160, allele-preferential binding prediction pointed towards preferential binding of TFs to the A allele, which was opposite of what we observed in the EMSA (Fig. 2c) but consistent with the luciferase assay (Fig. 2f, g). The transcription factors with the largest fold change in predicted binding *P*-values (20-80-fold) were the Activator Protein 1 (AP-1) transcription factors (Jun and Fos) (Table 3 and Supplementary Table 1) with the risk allele (G) disrupting the AP-1 motif (Fig. 4a).

**Figure 4:**
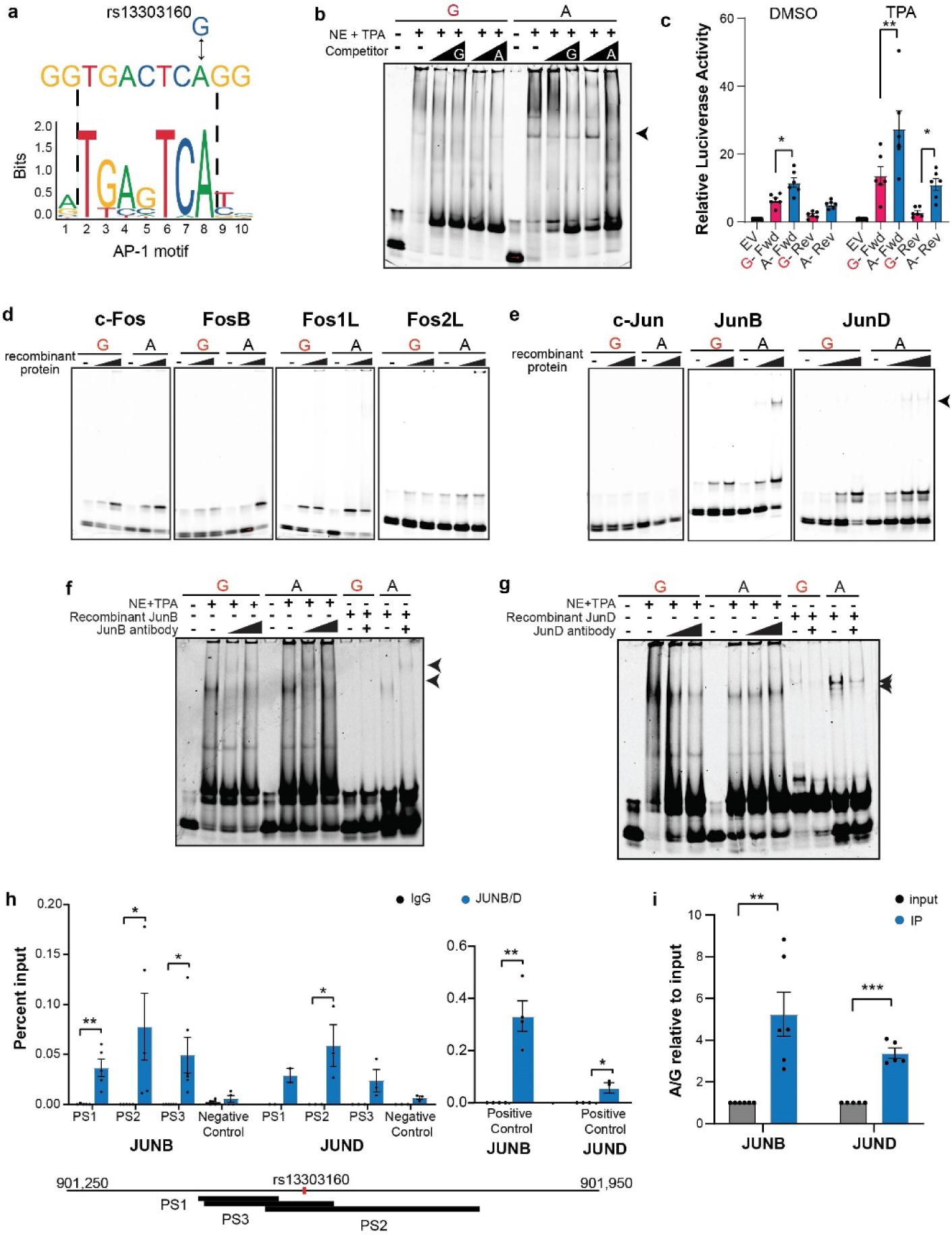
Allele-preferential binding of AP1 proteins to rs13303160 *in vitro* and *in vivo*. a) *in silico* TF binding predictions; b) EMSA using TPA-stimulated nuclear HeLa extract and fluorescently labeled oligonucleotide. Arrow indicates the allele-preferential binding; c) Luciferase reporter assay using DMSO and TPA stimulation and the rs13303160 sequence as an enhancer in the PANC-1 cell line; luciferase activity reported relative to the Empty Vector (EV). Unpaired t-tests were performed on the relative luciferase activity of the A/G ratio compared to A/A; d) Representative EMSAs with increasing amounts of recombinant Fos proteins (from left to right: c-Fos; FosB; Fos1L; Fos2L); e) Representative EMSAs with increasing amounts of recombinant Jun proteins (from left to right: c-Jun, JunB, JunD). Arrow indicates the allele-specific binding; f, g) Supershift EMSA with antibodies against JunB and JunD respectively using both TPA-stimulated nuclear lysate and recombinant protein; Arrows denote the shift in the bands; h) ChIP-qPCR in SW1990 PDAC cells for JunB and JunD using 3 primer sets (PS) surrounding the SNP. Positive controls are from a JunB ChIP-seq performed in the CFPAC1 PDAC cell line. Negative control is from a quiescent region on 1p36.33; i) TaqMan genotyping assay for rs13303160 using immunoprecipitated DNA from the ChIP. The ratio of A to G was determined relative to the quantity of A and G alleles in the input DNA. For all graphs, error bars represent the SEM. Unpaired two-tailed t-tests were performed; * *P* <0.05; ** *P* <0.01; *** *P* <0.001.

The binding motif for AP-1 (and sequence flanking rs13303160) is a TPA (12-O-Tetradecanoylphorbol-13-Acetate) response element indicating that TPA may induce expression and binding of AP-1 to these elements.^20^ We therefore repeated both the EMSA and luciferase experiments using cells treated with TPA. Upon TPA treatment, we observed an induction of AP-1 as demonstrated by the increase in JunB protein expression (Supplementary Fig. 3). EMSAs with TPA-treated HeLa nuclear extract demonstrated allele-preferential binding opposite what was originally seen (Fig. 2c) with preferential binding to the A allele over the G allele (Fig. 4b). In luciferase assays, cells treated with TPA demonstrated a stronger induction of enhancer activity in all three cell lines compared to vehicle control (Fig. 4c). However, the allele-preferential effects remained the same with the protective A allele showing higher activity as compared to the risk increasing G allele (FC 2-3.9, *P*=6.3×10^−3^-0.018). This indicates that AP-1 proteins may be responsible for the gene regulatory activity observed in the luciferase assays.

The AP-1 family of proteins includes the c-Fos and c-Jun proteins that can homo- or heterodimerize and play different roles in transcriptional regulation depending on the context.^20^ The *in silico* analysis did not predict which of the AP-1 protein family members bind rs13303160 as they all recognize the same motif. To determine which AP-1 protein(s) may exhibit allele-preferential binding *in vitro*, we performed EMSA using recombinant proteins for c-Fos, FosB, Fos-related antigen 1 (FRA-1), Fos-related antigen 2 (FRA-2) (Fig. 4d), Jun, JunB, and JunD (Fig. 4e). c-Fos and FosB demonstrated some allele-preferential binding, though inconsistently. Recombinant JunB and JunD proteins, on the other hand, demonstrated consistent allele-preferential binding. We subsequently performed supershift EMSAs with antibodies against JunB and JunD and observed a shift in the allele-preferential band when the antibody is added to the binding reaction indicating that the allele-preferential bands observed include JunB and JunD (Fig. 4f,g).

We then assessed if the *in vitro* allele-preferential binding of JunB and JunD translated to the context of genomic DNA. We performed ChIP-qPCR for JunB and JunD in the PDAC SW1990 and Hs766T cell lines (both heterozygous at rs13303160) and observed an enrichment of JunB with two primer sets that encompass the SNP and a third primer set just upstream of the SNP in SW1990 cells (Fig. 4h). We were unable to observe consistent enrichment in the Hs766T cell line with these primers. We additionally examined JunD localization at rs13303160 in the SW1990 cell line and observed a significant enrichment of JunD with primer set 2 (PS2) that encompasses the SNP (Fig. 4h). To determine if there was an allele-preferential enrichment in SW1990 cells, we quantified the immunoprecipitated DNA using a TaqMan genotyping assay. Compared to the input DNA, we observed an increased enrichment of the A allele over the G allele for both JunB (FC =5.2; *P*= 2.4×10^−3^) and JunD (FC=3.4 *P*=1×10^−5^) (Fig. 4i).

In summary, functional characterization of the seven fine-mapped SNPs led to the identification of three variants with allele-preferential binding *in vitro*, two of which also displayed allele-preferential gene regulatory activity. Transcription factor binding predictions and *in vitro* binding assays identified ELF2 and JunB/JunD to be mediating the allele-preferential effect at rs13303327 and rs13303160, respectively. Further *in vivo* analysis using ChIP-qPCR highlighted allele-preferential binding of JunB/D at rs13303160. Thus, we conclude rs13303160 represents a functional variant at the chr1p36.33 pancreatic cancer risk locus.

### Identification of likely target genes mediating the GWAS signal

Chr1p36.33 is gene dense with over 30 protein coding genes, microRNAs, and long non-coding RNAs. Using expression quantitative trait locus (eQTL) analysis, we identified two potential target genes at this locus for the tag SNP rs13303010 using the pancreas-specific GTEx (v8, n=305) dataset: *KLHL17* (Normalized Effect Size, NES = 0.35; *P* value = 4.9×10^−9^) and *NOC2L* (NES = −0.25; *P* value = 3.2×10^−5^) (Fig. 5a). Co-localization analysis^21^ of the GTEx (v7) pancreas eQTL and the PDAC GWAS signals indicated that the *KLHL17* and *NOC2L* eQTL may share a single causal variant with the GWAS signal (posterior probability = 0.99 and 0.75, respectively). Further, a transcriptome-wide association study (TWAS) from our group identified *KLHL17* as a borderline significant gene in the pancreas and multi-tissue models (Z = −3.96, FDR = 0.052; FDR = 0.063, respectively).^22^ Due to the stronger co-localization probability, suggestive TWAS results, and corroborating *in vitro* allele-preferential activity, we focused on *KLHL17* as a likely target gene underlying the chr1p36.33 risk signal.

**Figure 5:**
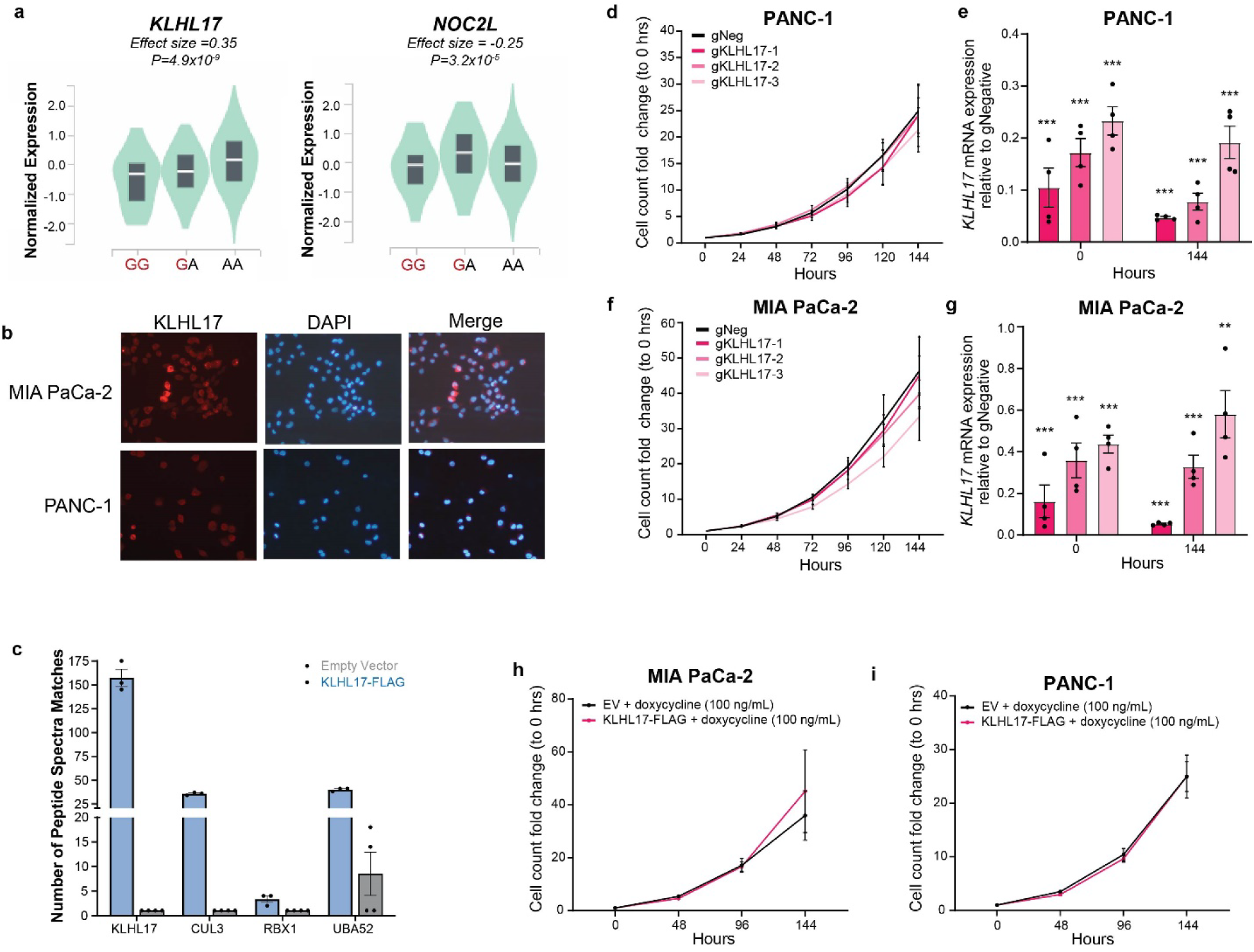
Analysis of the effects of altered KLHL17, a Cullin3-E3 complex member, expression on cellular growth of PDAC cells *in vitro*. a) Pancreas GTEx v8 eQTLs for rs13303010; b) Immunofluorescence for KLHL17 in the MIA PaCa-2 and PANC-1 KLHL17 overexpressing cell line; c) Peptide Spectra Matches for Cullin3-E3 members identified by KLHL17-FLAG immunoprecipitation and mass spectrometry analysis; d,f) Cell counts normalized to 0 Hours for CRISPRi-mediated knockdown of *KLHL17* in PANC-1 and MIA PaCa-2 cells, respectively (left panel). sgNeg is the negative control targeting a sequence within the same topologically associated domain (TAD). e,g) qPCR analysis of CRISPRi gRNA efficiency; expression is relative to the sgNeg control and internal HPRT control (right panel) and measured at the start and end of the growth assay; h, i) Cell counts normalized to 0 hours for doxycycline-inducible KLHL17-FLAG overexpressing and empty vector (EV) control MIA PaCa-2 and PANC-1 cell lines, respectively. For panels c-g, error bars indicate the SEM; For panels d-g, unpaired, two-tailed t-tests were performed, ** *P* <0.01, *** *P* <0.001

### Investigating the function of KLHL17 in the pancreas and in PDAC risk

#### KLHL17 is a part of the CULLIN3-RING ubiquitin ligase complex

KLHL17 is a member of the Kelch family of proteins. These proteins are substrate recognition proteins for the Cullin3-E3 ubiquitin complex,^23^ but the function of KLHL17 in the pancreas has not been defined. We therefore generated doxycycline-inducible KLHL17-FLAG tagged overexpressing PANC-1 and MIA PaCa-2 cell lines (Supplementary Fig. 4a) to study its function in the pancreas. We first examined KLHL17’s cellular localization using immunofluorescence. In the MIA PaCa-2 and PANC-1 overexpressing cell line, KLHL17 localized throughout the cell in contrast to previous reports indicating it is in the nucleoplasm and nuclear bodies^24^ (Fig. 5b). We then assessed whether KLHL17 is a member of the Cullin3-E3 ubiquitin complex. Using whole cell lysates from the PANC-1 induced overexpression and empty vector control cell lines, we performed an immunoprecipitation with a FLAG antibody (Supplementary Fig. 4b, c) followed by mass spectrometry to identify proteins interacting with KLHL17. We identified enrichment of KLHL17 along with CUL3, RBX1, and UBA52 in our KLHL17-FLAG IP but not the in the empty vector control IP (Fig. 5c) indicating that KLHL17 is a part of the CRL3 ubiquitin ligase complex in pancreatic cells.

#### KLHL17 overexpression and knockdown does not influence cell growth

We next assessed if KLHL17 plays a role in pancreas cell proliferation and cell viability. Knockdown of *KLHL17* using an siRNA pool in the PANC-1 and MIA PaCa-2 PDAC cell lines revealed a significant reduction in proliferation for both cell lines four days following knockdown (Supplementary Fig. 5a,c). However, we observed limited knockdown efficiency for *KLHL17* and a non-specific reduction of the nearby *NOC2L* gene (Supplementary Figure 5b, d). As these results did not distinguish whether the reduced levels of *KLHL17* or *NOC2L* mediated the growth effects observed, we implemented CRISPR interference (CRISRPi) in the PANC-1 and MIA PaCa-2 cell lines using guide RNAs targeting the 5’ untranslated region (UTR) of *KLHL17*. CRISPRi-mediated knockdown of *KLHL17* significantly reduced *KLHL17* mRNA levels in PANC-1 and MIA PaCa-2 cell lines (Figure 5e, g) while minimally affecting *NOC2L* expression (Supplementary Fig. 5e, f). Surprisingly, despite the significant reduction in *KLHL17* mRNA levels, KLHL17 protein levels remain unchanged even 36 days after transduction (Supplementary Fig. 5g, h). Subsequent growth analysis revealed that inhibition of *KLHL17* mRNA expression did not affect cell proliferation in the two PDAC cell lines (Fig. 5d, f). Furthermore, overexpression of *KLHL17* in PANC-1 and MIA PaCa-2 cells did not alter cell growth (Fig. 5h, i). We hypothesize that the initial growth phenotype observed with *KLHL17* siRNA is likely a result of the non-specific reduction in *NOC2L* expression as targeted knockdown of *NOC2L* with siRNA also reduces cell growth (Supplementary Fig. 5i-l).

#### KLHL17 may influence inflammatory pathways in the pancreas

Since KLHL17 did not have an evident role in PDAC cell growth, we sought to investigate potential functional consequences of altered *KLHL17* expression in the pancreas using an *in silico* knockdown analysis as previously described.^22,25^ We performed this analysis using the GTExv8 Pancreas RNA-seq dataset. Samples were separated into quartiles based on *KLHL17* gene expression and the upper (n=82) and lower (n=82) quartiles (Supplementary Fig. 6a) were subjected to differential gene expression analysis with EdgeR.^26^ We then used Gene Set Enrichment Analysis (GSEA) and Ingenuity Pathway Analysis (IPA) on the significantly differentially expressed genes (GSEA: n=13,090, FDR <0.05; IPA: n=3,511, FDR <0.05 and log2 fold change > |0.5] (FC=1.4)) to identify enriched pathways. We observed a positive enrichment of gene sets involved in inflammation or inflammation-related diseases (Fig. 6a-c). Further, IPA indicated similar results with nine of the ten most significant pathways being related to inflammation (Supplementary Fig. 6b) and eight of the nine pathways predicted to be activated in the GTEx samples with lower *KLHL17* expression. This indicates that lower KLHL17 expression may be associated with a pro-inflammatory environment in the pancreas.

**Figure 6:**
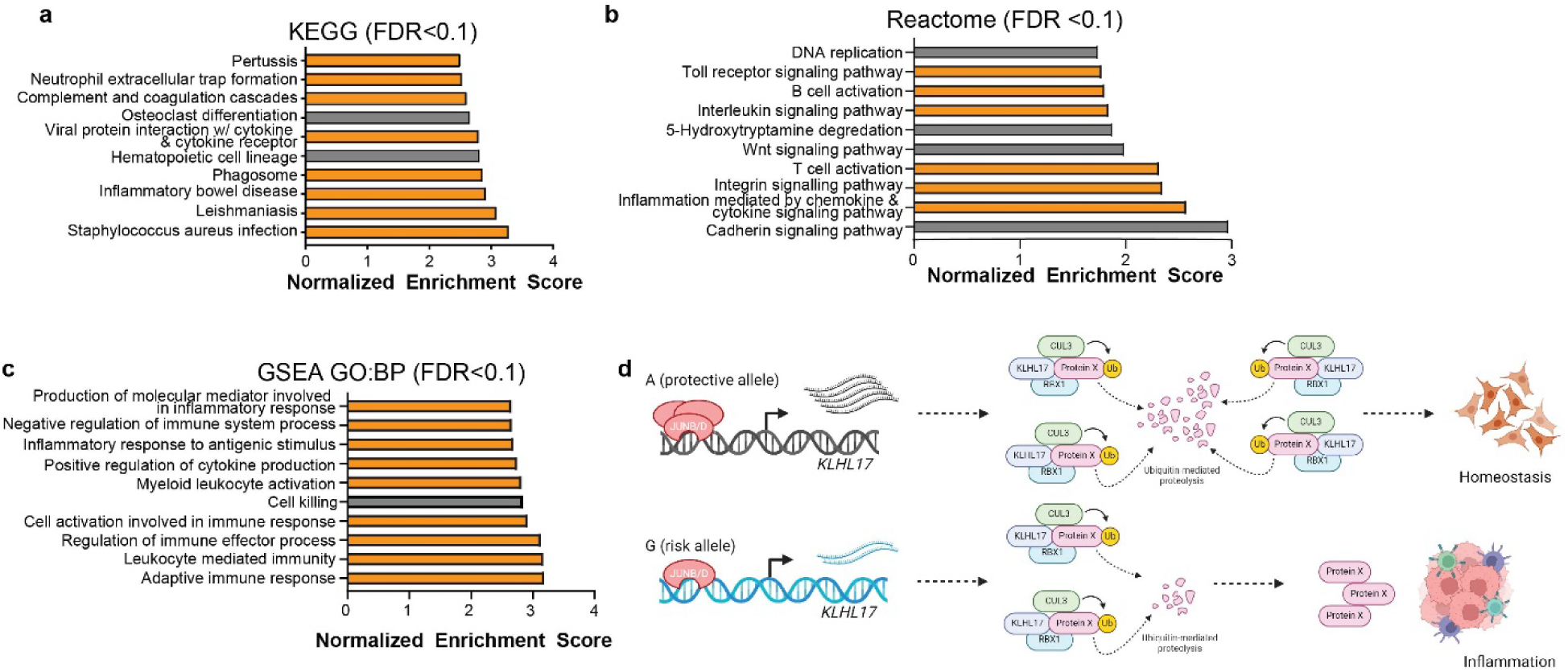
Gene Set Enrichment Analysis (GSEA) of differentially expressed genes from an *in silico KLHL17* knockdown. a) GSEA using the KEGG functional database and the significantly (FDR < 0.05) differentially expressed genes when KLHL17 expression is lower; b) GSEA using the Reactome dataset; c) GSEA using Biological Processes Gene Ontology dataset. For all three analyses only gene sets with an FDR < 0.1 are shown. Bars in orange indicate gene sets associated with inflammation or inflammation-related diseases. No gene sets were negatively enriched at this FDR threshold; d) Working hypothesis for the function of rs13303160 and KLHL17 in PDAC risk

We hypothesize that KLHL17 plays a role in mitigating inflammation by recruiting pro-inflammatory proteins for ubiquitination and degradation. This suggests that lower expression of KLHL17 that is associated with risk promotes a pro-inflammatory environment that is prime for tumorigenesis (Fig. 6d).

## Discussion

Here, we functionally characterize a common PDAC risk locus at chr1p36.33. Fine mapping identified a set of seven candidate functional variants for further analysis. *In vitro* binding assays narrowed this set to three SNPs, namely rs13303010, rs13303327, and rs13303160 that exhibited allele-preferential protein binding. Subsequent luciferase experiments revealed *in vitro* allele-preferential gene regulatory activity for rs13303327 and rs13303160, but not rs13303010. Using *in silico* prediction and *in vitro* binding studies, we identified ELF2 and JunB/D as transcription factors mediating the binding preference to the protective alleles of rs13303327 and rs13303160, respectively. This binding preference for JunB and JunD at rs13303160 was confirmed in the context of native chromatin using ChIP-qPCR followed by TaqMan genotyping assays. Taken together, we conclude that rs13303160 is a functional SNP at 1p36.33 whose effect is mediated through allele-preferential binding of JunB and JunD.

eQTL analysis in GTEx pancreas samples identified two possible target genes, *KLHL17* and *NOC2L*. *NOC2L* encodes the protein Novel INHAT Repressor which is known to repress p53 and histone acetyltransferase activity.^27^ *KLHL17* encodes for Actinfilin, a known substrate recognition protein for the Cullin3-RING ligases (CRL3), in neurons.^28^ We focused on characterizing *KLHL17* as a functional gene underlying the 1p36.33 GWAS signal for multiple reasons: 1) there is strong co-localization of the *KLHL17* eQTL with the GWAS signal; 2) *KLHL17* is a suggestive PDAC TWAS gene;^22^ and 3) the allele-preferential luciferase regulatory activity and TF binding were congruent with the *KLHL17* eQTL.

The KLHL family, comprised of 42 KLHL proteins, has been reported to have countless roles in cancer including gastrointestinal cancers.^23,29^ Canonically, KLHL proteins are substrate adaptor proteins for CRL3, an E3 ubiquitin ligase^30^ that are responsible for mediating the recognition, ubiquitination and degradation of their protein substrate(s) and are involved in a variety of cellular processes.^23^ Changes in KLHL protein expression affect substrate protein expression and downstream pathways.^31,32^ Depending on the KLHL family member and the cancer context, both increased and decreased KLHL family member expression has been described in the context of carcinogenesis.^23,29^ Recently, there has been an interest in targeting KLHL proteins to uncover their mechanisms and as a therapeutic strategy for associated diseases such as cancer.^33^

Only one study has characterized KLHL17’s role in cancer.^34^ In Liu et al., the authors highlighted a non-canonical role for *KLHL17* in the Ras/Map Kinase (MAPK) pathway where *KLHL17* overexpression enhances cell proliferation, migration, and colony forming ability for non-small cell lung cancer (NSCLC).^34^ They did not indicate if the involvement with MAPK could be through protein ubiquitination, as is known for the KLHL proteins and KLHL17 in neurons.^28^ In PDAC-derived cells, we observed that KLHL17 associates with members of the CRL3 family supporting a canonical function for this protein in the pancreas, as a CRL substrate recognition protein.

Further, Liu and colleagues reported an overexpression of *KLHL17* in NSCLC tumors compared to adjacent-normal tissue from TCGA.^34^ In the pancreas, there was no significant difference in *KLHL17* expression between TCGA PAAD tumor samples and normal or normal-adjacent tissue.^35^ However, eQTL and TWAS indicate that lower *KLHL17* expression levels associate with increased PDAC risk.^22,36^ We sought to characterize KLHL17’s function on pancreas cell growth but were presented with several technical challenges: 1) our initial knockdown experiments using siRNA against *KLHL17* revealed a growth suppression, but knockdown efficiency was minimal and non-specific; 2) CRISPRi-mediated knockdown of *KLHL17,* which did not demonstrate a growth phenotype, drastically reduced *KLHL17* mRNA expression, however, protein levels remained unchanged, even after 36 days. This confounding finding means that we cannot rule out a growth phenotype for different KLHL17 protein levels. We hypothesize that the initial growth phenotype observed with *KLHL17* siRNA can be contributed to the non-specific knockdown of *NOC2L,* as specific knockdown of *NOC2L* using siRNA also suppressed cell growth. This suggests that *NOC2L* may play a role in PDAC tumorigenesis.

Since our functional analysis pointed to *KLHL17* as a functional gene underlying the chr1p36.33 risk signal, we utilized an agnostic approach to identify the function of KLHL17 in the pancreas. *In silico* differential gene expression analysis using GTEx pancreas samples highlighted an enrichment of upregulated genes involved in inflammation-related pathways and gene sets with lower KLHL17 expression. This suggests that lower levels of KLHL17 may be associated with a pro-inflammatory state in the pancreas, or decreased ability to resolve the consequences of inflammatory signals.

Inflammation is known to be a contributing risk factor to the development of PDAC.^37^ PDAC arises from pancreatic intraepithelial neoplasia (PanIN) that display cancerous and pancreatic duct cell properties. Although PanIN exhibit duct-like properties, multiple lines of evidence indicate that pancreatic acinar cells that have undergone acinar-to-ductal metaplasia (ADM) are precursors for PDAC.^38^ ADM is a trans-differentiation process in which acinar cells lose acinar specific markers and gain duct cell markers. The plasticity of acinar cells makes them highly sensitive to external stimuli.^39^ Acinar cells can recover from an acute stimulus, but with a more sustained stimulus, such as during chronic inflammation, ADM can become irreversible resulting in progression to PanIN.^40^ Previous mouse studies have demonstrated that haploinsufficiency of *Nr5a2*, a likely GWAS risk gene and key acinar transcription factor, can impede pancreatic epithelial cells’ ability to recover from inflammation, resulting in increased ADM and PanINs.^25,41^ Unfortunately, our *in vitro* model system is limited with regards to understanding the effects of inflammation and ADM in the context of KLHL17 expression in the pancreas *in vivo*. Most pancreas cell lines are derived from tumors or metastatic lesions and have ductal characteristics. Available human normal-derived pancreatic cell lines are duct epithelial cells and would not recapitulate ADM under such stimuli. Primary human pancreas cells, particularly acinar cells, are difficult to maintain in culture as the acinar cell population is quickly lost due to ADM and cell death.^42^ Due to these limitations, additional studies are needed to investigate the role of KLHL17 in the context of inflammation, acinar cells and pancreatic tissue.

Taken together, we propose an explanatory model for the chr1p36.33 PDAC GWAS locus wherein lower KLHL17 expression allows for unresolved inflammation and acinar cell injury resulting in increased likelihood of the development of PanINs and progression to PDAC. Under cellular stress, JunB and JunD are induced by extracellular stimulus.^43^ Under such conditions, the preferential binding of JunB/D to the protective allele at rs13303160 may drive KLHL17 expression, resulting in the recognition of key inflammatory protein(s) for ubiquitination and degradation and thereby mitigate the inflammation. In contrast, when the risk allele is present, JunB/D may not sufficiently induce KLHL17 expression to resolve this inflammation (Fig. 6d). Further *in vivo* studies are needed to fully explore the role of KLHL17 in the pancreas especially in modulating carcinogenesis.

## Materials and Methods

### UK Biobank PDAC GWAS and meta-analysis

We obtained the cancer registry and hospital inpatient information phenotype data from UKBB on August 6^th^, 2021. Case criteria for patients diagnosed with pancreatic ductal adenocarcinoma (PDAC): [22006-0.0]=1 (white British), [22010-0.0]≠1 (recommended genomic analysis exclusions), [22019-0.0]≠1 (sex chromosome aneuploidy exclusion), [31-0.0] (self-reported sex; 0=female, 1=male) is consistent with [22001-0.0] (genetic sex), [40012] (Tumor behavior) include only: ‘3’ – malignant, primary site, [40008] (age at cancer diagnosis), [40006] – Type of Cancer ICD10 codes (C25* - except for C25.4). Control criteria: exclude control if any of the following Data Fields have values >0, [134] – Number of self-reported cancers, [2453] – Cancer diagnosed by doctor; exclude if any ICD10 codes in the following starting with C*, [40006] – Type of Cancer ICD10 codes, [41202] – Diagnoses – main ICD10 summary information, [41204] – Diagnoses – secondary ICD10 summary information, [41270] Diagnoses – ICD10 from Hospital inpatient information; exclude if any question in the following with a value that is not “NA”, [20001] - Cancer code self-reported, [20007] – Interpolated age when cancer first occurred, [40001] - Underlying primary cause of death ICD10; [40008] - Age at cancer diagnosis, [40011] - Histology of cancer tumor, [40012] - Behavior of cancer tumor, [40013] - Type of cancer ICD9 codes, [84] - Cancer year/age first occurred (Medical conditions), [40009] - Reported occurrences of cancer – Cancer Register; [31-0.0] (self-reported sex) is consistent with [22001-0.0] (genetic sex); [22010] - Recommended genomic analysis exclusions – filter samples based on poor heterozygosity/missingness as per UKB analysis (exclude if “1”); [22018] - Genetic Relatedness exclusions - exclude those with “1” or “2”; [22019] – Sex chromosome aneuploidy – exclude those with ‘Yes’; [22021] - Genetic kinship to other participants – only use those with “No kinship found”. We select up to 10 controls for each case based on similar age groups (same gender and birthyear +/− 5 years) and genetic background (using PCAmatchR^44^). The imputed genotype data was downloaded from UKBB (Nov. 2021). We removed the variants with MAF < 0.5%, INFO < 0.3, and complete rate > 10%. Genome-wide association test was performed by SNPTEST(v2.5.4-beta3)^45^ with covariates (age, gender, significant principal components and array type).

Summary statistics using a “Frequentist” (additive) model from four previously published GWAS phases (PanScan I-III)^5–8^ and PanC4,^9^ and the UK Biobank summary statistics described above (case subjects: n = 10,106 and control subjects: n = 21,895) were used for the meta-analysis. The meta-analysis was performed using Metal.^46^

All studies obtained consent from participants and Institutional Review Board (IRB) approvals including IRB certifications permitting data sharing in accordance with the NIH Policy for Sharing of Data Obtained in NIH Supported or Conducted Genome-Wide Association Studies (GWAS). Additionally, the PanScan study was approved by the NCI Special Studies Institutional Review Board.

### Fine-mapping of the GWAS signal

The chr1p36.33 GWAS region was fine-mapped using the chi-squared likelihood ratio test of the GWAS *P*-values for all SNPs. Linkage disequilibrium (LD) with the tag SNP rs13303010 was determined for every GWAS variant between chr1:1-2,300,000 (hg19) using LDLink^47^ European population. Variants with a LLR < 1:100 and an LD r^2^ > 0.8 were considered likely functional SNPs for experimental validation. Additionally, the Sum of Single Effects (SuSiE) linear regression, a Bayesian approach, was used to identify credible sets of variants likely harboring functional variant(s).^12^ A threshold of 0.9 probability that the credible set of variants contains a causal SNP and L=10 (up to 10 variants in a credible set) was used. PanScan III data (cases and controls) was used to generate the required LD matrix.^7,8^

### Electrophoretic Mobility Shift Assays (EMSA)

SNP-centered 31 bp oligonucleotides (Supplementary Table 2) were labeled with IRDye©700 fluorescent dye on the 5’ end and HPLC purified (Integrated DNA Technologies, Coralville, IA). Competitor oligonucleotides were unlabeled (IDT). Oligonucleotides were annealed at 99 °C and cooled slowly to room temperature. EMSA binding reactions were as follows: 10X binding buffer, 50% glycerol, polyDiDC, nuclear lysate (2.5-5 µg, ActiveMotif, Carlsbad, CA), labeled oligo (5nM), water. Competition binding reactions included unlabeled oligos at 50X and 100X labeled oligo concentration. Reactions were incubated at room temperature in the dark for 20 minutes. Reactions were then loaded on 4-12% gradient TBE gels (Invitrogen, Waltham, MA) with 0.5X TBE and ran for 100 min at 90V. Gels were imaged on the BioRad ChemiDoc^TM^ with the IR680 setting. For the TPA-stimulated EMSAs, two-hour TPA stimulated HeLa nuclear extract (ActiveMotif, Carlsbad, CA) was used.

For supershift assays, the nuclear lysate and antibodies for ELF2, JunB or JunD were incubated for 20 minutes at room temperature prior to the addition of other binding reaction components. The reactions were incubated for an additional 20 minutes.

For EMSAs with recombinant proteins, 135ng or 270ng of recombinant protein was used in place of nuclear lysate. Recombinant ELF1 (TP760629), ELF2 (TP760288), ELF3 (TP300631), ELF4 (TP761826), JUNB (TP303595), JUND (TP316958 4), c-FOS (TP760257), FOS1L (TP302104), FOSB (TP762032), FOS2L (TP760114) protein were purchased from Origene (Rockville, MD). c-JUN was purchased from Abcam (Waltham, MA) (ab84134). ELF5, which has negligible expression in the pancreas, was not tested.

### Plasmids

Luciferase backbone plasmids used for reporter assays were pGL4.23 and pGL4.14 (Promega, Madison, WI). Four gene blocks (forward orientation reference allele, forward orientation alternate allele, reverse orientation reference allele, reverse orientation alternate allele) of varying sizes (141-201 bps) were synthesized for rs13303160, rs13303010, and rs7524174 (Integrated DNA Technologies, Coralville, IA). Sequences to be assayed (Supplementary Table 3) were determined based on ATAC-seq and ChromHMM annotations previously generated in PDAC cell lines ^17,18^ with the goal of being inclusive of regulatory elements as well as sequence complexity. A 165 bp sequence for rs13303327 was cloned from a heterozygous HapMap CEPH subject (NA12716). The gene blocks were inserted into the luciferase plasmid as either enhancers or promoters. *KLHL17* pcDNA3 plasmid was purchased from GenScript (Piscataway, NJ) and subcloned into the pFUGW with a TREG3 promoter for tetracycline-inducible lentiviral expression. For CRISPRi experiments, stably expressing dCas9-KRAB-ZIM3 cell lines were generated using pLX303-ZIM3-KRAB-dCas9 (Addgene, Watertown, MA). Guide RNAs were purchased from Integrated DNA Technologies (Coralville, IA) and cloned into pU6-sgRNA EF1Alpha-puro-T2A-BFP (Addgene, Watertown, MA #60955). Guide RNA sequences (Supplementary Table 4) for CRISPRi were determined using the UCSC Genome Browser CRISPR targets track that uses the CRISPOR program.^48^ The negative targeting control (gNegative) is targeted to an open chromatin region within the same topologically associated domain as *KLHL17*. Sequences can be found in Supplementary Table 3. siRNA pools for *KLHL17*, *NOC2L*, and non-targeting were purchased from Horizon Discovery Dharmacon (Lafayette, CO).

### Cell Culture

Cell lines (MIA PaCa-2, PANC-1, SW1990, Hs766T, and HEK293T) were purchased from ATCC (Manassas, VA). PANC-1, Hs766T, and HEK293T cells were maintained in DMEM with 10% FBS. MIA PaCa-2 cells were maintained in DMEM with 2.5% horse serum and 10% FBS. SW1990 cells were maintained in RPMI and 10% FBS. All cells were grown at 37°C with 5% CO2. For virus production, HEK293T cells were grown in high glucose DMEM media supplemented with 10% FBS, 1% glutamine, and 1% sodium pyruvate. Cell lines were routinely tested for mycoplasma and were always found to be negative. Cell lines were also tested for authentication with a panel of short tandem repeats (STRs) via the Identifiler kit (Life Technologies, Carlsbad, CA) and compared with ATCC and DSMZ (German Collection of Microorganisms and Cell Cultures – https://www.dsmz.de/) STR profile datasets. All cell lines with profiles in the databases matched and those not with profiles in this database matched earlier passages of these cell lines in use in our laboratory.

### Lentivirus production

HEK293T cells were plated 24 hours prior to transfection. The plasmid of interest and packaging vectors (x2, MDG2) were transfected using Lipofectamine 3000 (Thermo Fisher Scientific, Waltham, MA) and media was changed 6-8 hours post-transfection. Forty-eight hours later, virus was harvested, filtered with a 0.45-micron syringe filter, and precipitated overnight at 4°C with 2X PEG precipitation buffer. Precipitated virus was centrifuged at 1000xg for 30 min, 4°C, the supernatant was removed and pelleted virus resuspended in PBS.

### Generation of stable cell lines

MIA PaCa-2 and PANC-1 cell lines were used to make stably expressing dCas9-KRAB-Zim3 and KLHL17 doxycycline-inducible KLHL17-FLAGoverexpressing lines. For stably expressing dCas9 lines, cells were transduced with virus for 24 hours. After 24 hours, selection with 10µg/mL of blasticidin was initiated. Once selection was complete, cells were diluted in a 96 well plate at a seeding density of 0.5 cells/well to isolate individual cell colonies. These colonies were expanded and protein expression was verified by western blot. Transduction with CRISPRi gRNAs was performed in the same manner with the stably expressing dCas9 and selected with 8-10µg/mL of puromycin. To generate doxycycline-inducible KLHL17-FLAG cell lines, previously generated MIA PaCa-2 and PANC-1 cells stably transduced with the Clontech (Mountain View, CA) TetOn3G transactivator plasmid (pLVX-Tet3G),^49^ were transduced with the TREG3p-FUGW-KLHL17 lentiviral expression plasmid (described above) in a 12-well plate. Media was changed 24 hours post-transduction and puromycin selection (8-10 µg/mL) initiated at 48 hours.

### Luciferase Assays

MIA PaCa-2, PANC-1 and HEK293T cells were seeded in 48-well plates 24 hours prior to transfection. Cells were co-transfected with 1 µg of the luciferase plasmid pGL4.14 or pGL4.23, (Promega, Madison, WI) and 35 ng of pGL4.74 Renilla vector (Promega, Madison, WI) using Lipofectamine 2000 (Thermo Fisher Scientific, Waltham, MA). Forty-eight hours following transfections, cells were washed twice with PBS. Luciferase assays were performed with the Promega Dual Luciferase Reporter Assay following the manufacturer’s instructions. Luciferase activity was normalized to Renilla luciferase activity and reported as the fold-change relative to the empty luciferase vector. A Student’s two-tailed t-test was performed to test for statistically significant differences between alleles. For luciferase assays with TPA, cells were simultaneously transfected and stimulated with 200nM Phorbol 12-myristate 13-acetate (TPA, Millipore Sigma, Burlington, MA) or DMSO 24 hours after plating and harvested 48 hours after transfection. Luciferase activity was determined as described above. Significance testing was performed on the ratio of A to G relative luciferase activity using a two-tailed t-test.

### Cell growth assays

For *KLHL17* overexpression cell proliferation assays, overexpression in stable cell lines was induced with 100 ng/mL of doxycycline 24 hours prior to plating. Cells were then plated in 12 well plates. Twenty-four hours after plating (defined as 0 hours), cell images were taken on the Lionheart (Biomek, Brea, CA) plate reader every 48 hours for seven days.

For *KLHL17* and *NOC2L* knockdown cell growth assays, cells were plated 24 hours prior to transfection in a 6 well. Cells were then transfected with siRNA pools against *NOC2L*, *KLHL17*, non-targeting control (Horizon Discovery Dharmacon Lafayette, CO) using RNAiMAX (Thermo Fisher Scientific Waltham, MA). Cell counts were then taken using the Lionheart (0 hour). Counts were taken daily for 7 days. At 72 hours, cells were re-transfected with siRNA. For CRISPRi growth assays, cells were plated in a 12-well plate following complete selection with puromycin. The first count was taken 24 hours after plating and considered 0 hours. Counts were subsequently taken every 24 hours for 7 days. For all cell proliferation assays, the cell count for each day was normalized to the 0-hour cell count and is represented as a fold change relative to the initial cell count.

### RNA isolation and Reverse Transcriptase quantitative PCR

RNA was isolated using the QIAGEN RNeasy kit with a DNase digest and the QIAcube (QIAGEN, Germantown, MD). RNA was reverse transcribed to cDNA using SuperScript III Reverse Transcriptase (Invitrogen, Waltham, MA). Gene expression levels were quantified by qRT-PCR using TaqMan (Thermo Fisher Scientific, Waltham, MA) assays: *NOC2L* (Hs00610834_g1), *KLHL17* (Hs00938625_g1), *HPRT* (Hs99999909_m1).

### Chromatin Immunoprecipitation

Chromatin Immunoprecipitation was performed using the ActiveMotif (Calrsbad, CA) High Sensitivity ChIP kit with SW1990 and Hs766T PDAC cell lines. For JunB/D ChIP-qPCR, cell media was replaced approximately 16 hours prior with serum free media and cells were stimulated with 200nM of TPA (Millipore Sigma, Burlington, MA) for 3.5 hours prior to cross-linking. Following crosslinking, cells were lysed with a 25-gauge syringe prior to sonication. Samples were sonicated using the Covaris ME220 (Woburn, MA). Shearing efficiency and chromatin concentration was assessed for the input. Immunoprecipitation was set up following the ActiveMotif protocol. Two hundred microliters of sheared chromatin were used for IPs with either ELF2 (4µg), JunD (4µg), JunB (10µL) or IgG (4µg) antibodies. IPs were incubated at 4°C overnight, then protein G agarose beads were added for 3 hours. Following DNA purification, enrichment of ELF2, JunB, or JunD at the regions of interest was assessed using qPCR (Supplementary table 5 for primers) using SYBR Green Master Mix. Percent input was calculated following ActiveMotif’s protocol using a standard curve from input DNA for each primer set. Allelic enrichment was determined using TaqMan genotyping assays (C 57466801_10 and custom design). The ratio of A to G alleles was calculated and compared to the input DNA allelic ratio.

### Antibodies

Antibodies for EMSA: ELF2 (ab28726, Abcam; 12499-1-AP, Proteintech Rosemont, IL), JunB (C37F9, Cell Signaling Technologies, Danvers, MA), JunD (D17G2, Cell Signaling Technologies, Danver, MA). For Western Blot and Immunoprecipitation: JunB (1:1000, C37F9, Cell Signaling Technologies, Danver, MA), JunD (1:1000, D17G2, Cell Signaling Technologies, Danver, MA), FLAG (1:1000, F1804, Millipore Sigma, Burlington, MA), KLHL17 (1:500, PA5-56689 Thermo Fisher Scientific, Waltham, MA); 1:500, HPA031251, Millipore Sigma), GAPDH (1:1000, ab125247, Abcam, Waltham, MA), SP1 (1:000 ab13370, Abcam); mouse anti-rabbit light chain specific antibody HRP (1:5000, C840Z39 Jackson ImmunoResearch, West Grove, PA); donkey anti-mouse secondary HRP (1:5000, ab7061, Abcam, Waltham, MA), donkey anti-rabbit secondary HRP (1:5000, ab205722, Abcam, Waltham, MA); For ChIP: ELF2 (12499-1-AP, Proteintech, Rosemont, IL), JunB (C37F9,Cell Signaling Technologies, Danver, MA), JunD (720035, Invitrogen, Waltham, MA), Rabbit IgG (2729S, Cell Signaling Technologies, Danver, MA). Immunofluorescence: KLHL17 (HPA031251, Millipore Sigma, Burlington, MA), AlexaFluor647 (1:1000, A-31573, Thermo Fisher Scientific, Waltham, MA)

### Western blot analysis

Cells were lysed with either NP-40-DOC-SDS lysis buffer (150mM NaCl, 50mM Tris, 1% NP-40, 1% Sodium Deoxy cholate, 1% Sodium dodecyl sulfate) or NE-PER^TM^ Nuclear and Cytoplasmic extraction kit (Thermo Fisher Scientific, Waltham, MA). Lysates were run on Criterion^TM^ XT precast 3-8% Tris-Acetate gels (Bio-Rad, Hercules, CA) using XT running buffer and transferred to a PDVF membrane using a standard wet transfer or Bolt^TM^ 4-12% Bis-Tris Plus gels (Invitrogen, Waltham, MA) using MOPS running buffer and transferred to PDVF membranes using the iBlot^TM^ Transfer Stacks (Invitrogen, Waltham, MA). Membranes were blocked in 5% Bovine Albumin Serum and incubated with primary antibody overnight at 4°C. The appropriate HRP secondary antibody was added for a 1hr incubation at room temperature. Following washes with TBS-t, chemiluminescence was detected with SuperSignal™ West Femto Maximum Sensitivity Substrate (Thermo Fisher Scientific, Waltham, MA) and imaged on the Bio-Rad ChemiDoc^TM^.

### Immunoprecipitation

Cells were treated with 1µM of MG-132 proteosome inhibitor (Millipore Sigma, Burlington, MA)16 hours prior to harvest. Cells were harvested and lysed using 50 mM HEPES (pH 7.4), 150 mM NaCl, 0.5 mM EDTA, 0.1% NP-40, protease inhibitor, and 10µM MG-132 on ice for five minutes then freeze-thawed once. Samples were centrifuged at 14,000 rpm for 5 minutes to pellet cell debris. One milligram of protein extract was incubated with 2µg of FLAG antibody at 4^°^C for two hours. Protein G beads (Invitrogen, Waltham, MA) were added and incubated for an additional 30 minutes. Five IPs (for a total of 5mg of protein used) were combined and washed three times with 50mM HEPES. Ten percent of the IP was used for western blot analysis and the remaining 90% was subjected to mass spectrometry.

### Protein Digestion and TMT labeling

The cell pellets were lysed in EasyPrep Lysis buffer (Thermo Fisher, CA) according to manufacturer’s protocol. Lysates were clarified by centrifugation and protein concentration was quantified using BCA protein estimation kit (Thermo Fisher, CA). Fifteen micrograms of lysate were reduced, alkylated and digested by addition of trypsin at a ratio of 1:50 (Promega) and incubating overnight at 37°C.

For TMT labeling 100μg of TMTpro label (Thermo Fisher, CA) in 100% ACN was added to each sample. After incubating the mixture for 1 hr at room temperature with occasional mixing, the reaction was terminated by adding 50 μl of 5% hydroxylamine, 20% formic acid. The peptide samples for each condition were pooled and peptide clean-up was performed using the proprietary peptide clean up columns from the EasyPEP Mini MS Sample Prep kit (Thermo Fisher, CA).

### High pH reverse phase fractionation

The first dimensional separation of the peptides was performed using a Waters Acquity UPLC system coupled with a fluorescence detector (Waters, Milford, MA) using a 150mm x 3.0mm Xbridge Peptide BEM^TM^ 2. 5 μm C18 column (Waters, MA) operating at 0.35 ml/min. The dried peptides were reconstituted in 100 ul of mobile phase A solvent (10 mM Ammonium Formate, pH 9.4). Mobile phase B was 10 mM Ammonium Formate /90% acetonitrile, pH9.4. The column was washed with mobile phase A for 5 min followed by gradient elution 10-50% B (5-60 min) and 50-75 %B (60-70 min). The fractions were collected every minute. These 60 fractions were pooled into 24 fractions. The fractions were vacuum centrifuged to dryness and stored at −80°C until analysis by mass spectrometry.

### Mass Spectrometry acquisition and data analysis

The dried peptide fractions were reconstituted in 0.1%TFA and subjected to nanoflow liquid chromatography (Thermo Easy nLC 1200, Thermo Scientific, Thermo Scientific) coupled to an Orbitrap LUMOS mass spectrometer (Thermo Scientific, CA). Peptides were separated using a low pH gradient using a 5-50% ACN over 120 minutes in mobile phase containing 0.1% formic acid at 300 nl/min flow rate. MS scans were performed in the Orbitrap analyser at a resolution of 120,000 with an ion accumulation target set at 4e^5^ and max IT set at 50ms over a mass range of 400-1600 m/z. Ions with determined charge states between 2 and 5 were selected for MS2 scans. A cycle time of 3 sec was used and a quadrupole isolation window of 0.7 m/z was used for MS/MS analysis. An Orbitrap at 50,000 resolutions with a normalized AGC set at 250 followed by maximum injection time set as “Auto” with a normalized collision energy setting of 38 was used for MS/MS analysis.

Acquired MS/MS spectra were searched against a human uniprot protein database using a SEQUEST HT and percolator validator algorithms in the Proteome Discoverer 2.4 software (Thermo Scientific, CA). The precursor ion tolerance was set at 10 ppm and the fragment ions tolerance was set at 0.02 Da along with methionine oxidation included as dynamic modification. Carbamidomethylation of cysteine residues and TMT16 plex (304.2071Da) was set as a static modification of lysine and the N-termini of the peptide. Trypsin was specified as the proteolytic enzyme, with up to 2 missed cleavage sites allowed. Searches used a reverse sequence decoy strategy to control for the false peptide discovery and identifications were validated using percolator software. Only peptides with less the 50% co-isolation interference were used for quantitative analysis.

Reporter ion intensities were adjusted to correct for the impurities according to the manufacturer’s specification and the abundances of the proteins were quantified using the summation of the reporter ions for all identified peptides. The reporter abundances were normalized across all the channels to account for equal peptide loading. Data analysis and visualization were performed in Microsoft Excel or R.

### On-bead trypsin digestion and LC-MS/MS analysis

Beads were resuspended in 30 μL of 50 mM HEPES (pH 8.0) (pH 8.0) and heated at 95°C for 10 min. Samples were treated with 2 μg of trypsin and incubated at 37°C overnight with constant shaking. The supernatant containing the tryptic digests was collected after centrifugation. The residual beads were washed twice with 50 mM HEPES (pH 8.0), pH 8.0 and the supernatant and washes combined for maximum recovery. Peptides were desalted using EasyPep MS sample prep kit (Thermo Scientific, CA) and lyophilized. The dried peptides were suspended in 15 µL of 0.1% TFA and analyzed using an EASY-nLC 1200 (ThermoFisher Scientific, Waltham, MA) in front of an Q Exactive HF (ThermoFisher Scientific, Waltham, MA) equipped with an EasySpray ion source. The desalted tryptic peptide was loaded onto an Acclaim PepMap 100 (75 µM x 2 cm) C18 trap column (ThermoFisher Scientific, Waltham, MA) followed by a separation on PepMap RSLC C18 (75 µM x 25 cm) analytical column. The peptides were eluted with a 5% to 27% gradient of Acetonitrile with 0.1% Formic acid over 60 minutes and 27% to 40% gradient of Acetonitrile with 0.1% Formic acid over 45 minutes with a flow rate of 300 nl/min. The MS1 was performed at 60,000 resolutions over mass range of 380 to 1580 m/z, with a maximum injection time of 120 ms and an AGC target of 3e6. The MS2 scans was performed at resolution of 15,000, normalized collision energy set at 27, maximum injection time of 50 ms and an AGC target of 2e5.

MS files were searched with Proteome Discoverer 2.4 using the Sequest node. Data were searched against the Uniprot human database using a full tryptic digest, 2 max missed cleavages, minimum peptide length of 6 amino acids and maximum peptide length of 40 amino acids, an MS1 mass tolerance of 10 ppm, MS2 mass tolerance of 0.02 Da

### Immunofluorescence analysis

MIA PaCa-2 and PANC-1 cells overexpressing *KLHL17* were plated on coverslips and induced with 100ng/mL of doxycycline for 72 hours. Cells were then washed with PBS and fixed with 4% paraformaldehyde (Thermo Fisher Scientific, Waltham, MA) for 15 minutes. Following fixation cells were permeabilized with 0.25% Triton-X for 15 minutes. Samples were then blocked in 2% BSA (Millipore Sigma, Burlington, MA) for one hour at room temperature. Primary KLHL17 antibody was added to the cells at 1µg/mL and incubated at 4°C overnight. Cells were washed with PBS and incubated with an AlexaFluor 647 secondary antibody for one hour at room temperature in the dark. Coverslips were then mounted on slides using ProLong^TM^ Diamond Antifade Mountant with DAPI (Thermo Fisher Scientific, Waltham, MA) and allowed to cure for at least 24 hours at 4°C in the dark. Slides were imaged with Zeiss Microscope.

### In silico knockdown and pathway analysis

An *in silico* knockdown analysis for *KLHL17* was performed using GTExv8 normal pancreas tissue sample derived RNA-seq data as previously described.^22^ Briefly, we scaled GTExv8 pancreas gene expression counts to account for sequencing depth and RNA composition across all samples (n=328) to give normalized counts of the trimmed mean of M-values (TMM) using EdgeR.^26^ Genes with no reads for > 20% of the samples were excluded. Normalized reads were used to segregate samples into quartiles based on *KLHL17* expression. Only the samples in the top and bottom quartiles (n=82 each) were used for downstream analysis. The raw counts for these selected samples were re-normalized for sequencing depth to obtain pseudo-counts which were analyzed using the quantile-adjusted conditional maximum likelihood (qCML) method in EdgeR. Differential expression (log2[bottom/top quartile] and *P* values) was then assessed using an exact test. The statistically significant (FDR<0.05) differentially expressed genes were subjected to Gene Set Enrichment Analysis (GSEA) using webgestalt.org.^50^ The ranked list for the GSEA was based on log2 fold change. Without an FDR filter, there was no significant enrichment of gene sets. For Ingenuity Pathway Analysis, a FDR < 0.05 and a log2 fold change > |0.5| was used to filter genes for input (IPA, QIAGEN, Germantown, MD). For IPA, both the log2 fold change and FDR were considered in the analysis of enriched pathways.

### Transcription Factor Binding Prediction

In silico Transcription Factor binding prediction was performed using PrEdict Regulatory Functional Effect of SNPs by Approximate *P* value Estimation (PERFECTOS-APE; https://opera.autosome.org/perfectosape/). Briefly, the SNPs with allele-specific activity were submitted for analysis to determine the probability of a TF binding site from the position matrices in the HOCOMOCO11 transcription factor database. Once a *P*-value of predicted TF binding sequence was determined for each allele, a fold change was calculated.^19^

## Data Availability

The GWAS data is available on dbGAP and data in the paper are available upon reasonable request to the authors. UK Biobank data is available through the UK Biobank.

https://www.ncbi.nlm.nih.gov/projects/gap/cgi-bin/study.cgi?study_id=phs000206.v6.p3

https://www.ncbi.nlm.nih.gov/projects/gap/cgi-bin/study.cgi?study_id=phs000648.v1.p1

http://www.ukbiobank.ac.uk/

## Acknowledgements

This study utilized the high-performance computational capabilities of the Biowulf Linux cluster at the NIH, Bethesda, MD, USA (http://biowulf.nih.gov). The authors would like to thank the Frederick National Cancer Research Laboratories for the generation of the inducible plasmids. The authors would like to thank participants and clinical coordinators participating in the Pancreatic Cancer Cohort Consortium, Pancreatic Cancer Case-Control Consortium and the UK Biobank for providing samples for the GWAS studies.

The authors acknowledge the research contributions of the Cancer Genomics Research Laboratory for their expertise, execution, and support of this research in the areas of project planning, wet laboratory processing of specimens, and bioinformatics analysis of generated data.

The data used for the analyses described in this manuscript were obtained from the GTEx Portal versions 7 and 8 pancreas data.

This research has been conducted using data from UK Biobank, a major biomedical database (www.ukbiobank.ac.uk).^11^

