## Supplementary Table 5 for "Allelic effects on *KLHL17* expression likely mediated by JunB/D underlie a PDAC GWAS signal at chr1p36.33"

| Primer Set | Forward 5' - 3' | Reverse 5' - 3' |
|---|---|---|
| **rs1303327_PS1** | GAAGGAAGGCGAGACCTAGG | TCCGAGAAGCCCCCCTAGG |
| **rs13303327_PS2** | AGAAGGAAGGCGAGACCTA | CGTTGGACGCGGATTCTT |
| **rs13303327_PS3** | GGGTCCCATTCGACTTCTTG | TAGGTCTCGCCTTCCTTCTT |
| **ELF2 Positive Control (PYGO2)** | AGGCGTAGCGTCTCGTCCG | GAGCTGCAGCAACCACAAAGTG |
| **ELF2 Positive Control (TBP)** | GTGACCTATGCTCACACTTCTC | GAGTACAATCTGTTACCTGGGTC |
| **rs13303160_PS1** | GAGTCCCCTTAAGCCTTGGG | GAGTTCCAGCACAGGAGACG |
| **rs13303160_PS2** | TCTCCTGTGCTGGAACTCCT | CCTTTCTGGAAGAGGCCTGG |
| **rs13303160_PS3** | TTAAGCCTTGGGGACCCTGA | GCCCACACATCCTGTTTCCA |
| **Positive Control** | ATGCACGAGGCCTTTGAGAA | CAGGCAGTTCCTGTTGCCTA |
| **Negative Control** | GAACTCGAAAGGCACCAGGA | AAGGCCCTCTGGATGGATCT |

Supplementary Table 5: Sequences of primers used for ChIP-qPCR. Controls for ELF2 came from an ENCODE K562 GFP-ELF2 ChIP-seq. JunB/D positive control was determined from a JunB ChIP-seq in CFPAC1 cells (GSE119930). The negative control is from a quiescent region on chromosome 1 (4176991-4180055 (hg19))
