## Supplementary Table 4 for "Allelic effects on *KLHL17* expression likely mediated by JunB/D underlie a PDAC GWAS signal at chr1p36.33"

| Guide Name | Forward Strand (5'-3') | Reverse Complement (5'-3') |
|---|---|---|
| **gKLHL17-1** | TTGACGGACGCGGAGACTGCCGGGTTTAAGAGC | TTAGCTCTTAAACCCGGCAGTCTCCGCGTCCGTCAACAAG |
| **gKLHL17-2** | TTGCTCCGCGTCCGTTAAGCCCGGTTTAAGAGC | TTAGCTCTTAAACCGGGCTTAACGGACGCGGAGCAACAAG |
| **gKLHL17-3** | TTGGTCCTCCGCGAATCGGCGGTGTTTAAGAGC | TTAGCTCTTAAACACCGCCGATTCGCGGAGGACCAACAAG |
| **gNegative** | TTGTGGCTAGCAACATCTCGACAGTTTAAGAGC | TTAGCTCTTAAACTGTCGAGATGTTGCTAGCCACAACAAG |

Supplementary Table 4: Sequences for the gRNAs used in CRISPRi experiments. Guide sequences were determined using the CRISPOR feature on the UCSC Genome Browser.
