## Supplementary Table 3 for "Allelic effects on *KLHL17* expression likely mediated by JunB/D underlie a PDAC GWAS signal at chr1p36.33"

| | gBlock Sequences 5' - 3' |
|---|---|
| **rs13303010 Forward Ref** | TCA GCTAGC GCAGGAGTCACAGCTGCCCGCACGCCCAGCTCGCCCCAGCCCCGCTGAGAGGAGCAAGAAAAGCCCCCTTGGATA<br>CAGACACCCACCGGGAGGCCAAATCGGCCCTCGGACCCGCGGCTTACCTCTTGCGGCTCCCCGCAGCTGCCATGACACCAACCCG<br>AAGCGTGCACCCCACTTCCGGCCCCAGAATGCCGCGCGGCT AAGCTT CTG |
| **rs13303010 Forward Alt** | TCAGCTAGCGCAGGAGTCACAGCTGCCCGCACGCCCAGCTCGCCCCAGCCCCGCTGAGAGGAGCAAGAAAAGCCCCCTTGGATAC<br>AGACACCCACCGGGAGGCCAAATCAGCCCTCGGACCCGCGGCTTACCTCTTGCGGCTCCCCGCAGCTGCCATGACACCAACCCGA<br>AGCGTGCACCCCACTTCCGGCCCCAGAATGCCGCGCGGCT AAGCTTCTG |
| **rs13303010 Reverse Ref** | TCAGCTAGCAGCCGCGCGGCATTCTGGGGCCGGAAGTGGGGTGCACGCTTCGGGTTGGTGTCATGGCAGCTGCGGGGAGCCGCA<br>AGAGGTAAGCCGCGGGTCCGAGGGCCGATTTGGCCTCCCGGTGGGTGTCTGTATCCAAGGGGGCTTTTCTTGCTCCTCTCAGCGG<br>GGCTGGGGCGAGCTGGGCGTGCGGGCAGCTGTGACTCCTGC AAGCTTCTG |
| **rs13303010 Reverse Alt** | TCAGCTAGCAGCCGCGCGGCATTCTGGGGCCGGAAGTGGGGTGCACGCTTCGGGTTGGTGTCATGGCAGCTGCGGGGAGCCGCA<br>AGAGGTAAGCCGCGGGTCCGAGGGCTGATTTGGCCTCCCGGTGGGTGTCTGTATCCAAGGGGGCTTTTCTTGCTCCTCTCAGCGG<br>GGCTGGGGCGAGCTGGGCGTGCGGGCAGCTGTGACTCCTGC AAGCTTCTG |
| **rs13303160 Forward Ref** | TCAGCTAGCTGCGGTGGCTTTGGCCGCCGTCTCCTGTGCTGGAACTCCTGCCTCAGCCCTCCCTGCAGTCACCGGTGACTCGGGC<br>CGGCCAGAGTTTAGATGGAAACAGGATGTGTGGGCACGTTGTCCCGGGGGGCCTGGAAGGTCGCCCC GGAAGCTTCTG |
| **rs13303160 Forward Alt** | TCAGCTAGCTGCGGTGGCTTTGGCCGCCGTCTCCTGTGCTGGAACTCCTGCCTCAGCCCTCCCTGCAGTCACCGGTGACTCAGGC<br>CGGCCAGAGTTTAGATGGAAACAGGATGTGTGGGCACGTTGTCCCGGGGGGCCTGGAAGGTCGCCCC GGAAGCTTCTG |
| **rs13303160 Reverse Ref** | TCAGCTAGCGGGGCGACCTTCCAGGCCCCCCGGGACAACGTGCCCACACATCCTGTTTCCATCTAAACTCTGGCCGGCCCGAGTC<br>ACCGGTGACTGCAGGGAGGGCTGAGGCAGGAGTTCCAGCACAGGAGACGGCGGCCAAAGCCACCG AAGCTTCTG |
| **rs13303160 Reverse Alt** | TCAGCTAGCGGGGCGACCTTCCAGGCCCCCCGGGACAACGTGCCCACACATCCTGTTTCCATCTAAACTCTGGCCGGCCTGAGTC<br>ACCGGTGACTGCAGGGAGGGCTGAGGCAGGAGTTCCAGCACAGGAGACGGCGGCCAAAGCCACCG AAGCTTCTG |
| | Primers for amplification of rs13303327 from HapMap (165bp amplicon + vector overhang) |
| **rs13303327 Forward Orientation Forward Primer** | TCAGCTAGCGGAAGAAGGAAGGCGAGACCTAG |
| **rs13303327 Forward Orientation Reverse Primer** | CAGAAGCTTGCCTCGCCCTCCTCCTC |
| **rs13303327 Reverse Orientation Forward Primer** | CAGAAGCTTGGAAGAAGGAAGGCGAGACCTAG |
| **rs13303327 Reverse Orientation Reverse Primer** | TCAGCTAGCGCCTCGCCCTCCTCCTC |

Supplementary Table 3: Gene Block (gBlock) sequences used for luciferase assays. Assays were completed using the forward and reverse orientations of the sequence. Sequence lengths were determined based on the epigenomic annotations. For rs13303327, gBlocks could not be synthesized and was cloned from a HapMap subject heterozygous for the SNP using the primers listed
