## Supplementary Table 2 for "Allelic effects on *KLHL17* expression likely mediated by JunB/D underlie a PDAC GWAS signal at chr1p36.33"

| | Forward Oligo | Reverse Complement Oligo |
|---|---|---|
| **rs13303010 REF** | CCGGGAGGCCAAATC**G**GCCCTCGGA CCCGCG | CGCGGGTCCGAGGGC**C**GATTTGGCC TCCCGG |
| **rs13303010 ALT** | CCGGGAGGCCAAATC**A**GCCCTCGGA CCCGCG | CGCGGGTCCGAGGGC**T**GATTTGGCC TCCCGG |
| **rs13303160 REF** | AGTCACCGGTGACTC**G**GGCCGGCCA GAGTTT | AAACTCTGGCCGGCC**C**GAGTCACCG GTGACT |
| **rs133030160 ALT** | AGTCACCGGTGACTC**A**GGCCGGCCA GAGTTT | AAACTCTGGCCGGCC**T**GAGTCACCG GTGACT |
| **rs13303327 REF** | CTTCCCAGAGGAGGA**G**GATGGCGGG GCCTGG | CCAGGCCCCGCCATC**C**TCCTCCTCTG GGAAG |
| **rs13303327 ALT** | CTTCCCAGAGGAGGA**A**GATGGCGGG GCCTGG | CCAGGCCCCGCCATC**T**TCCTCCTCTG GGAAG |

Supplementary Table 2: Sequences of oligonucleotides used for EMSAs. The IRDye700 and unlabelled oligonucleotides are the same.
