## Supplementary Table 1 for "Allelic effects on *KLHL17* expression likely mediated by JunB/D underlie a PDAC GWAS signal at chr1p36.33"

Supplementary Table 1: In silico TF motif predictions for rs13303327 and rs13303160

| rsID | Motif | Allele 1 | Allele 2 | Allele 1 *P*-value | Allele 2 *P*-value | *P*-value Fold Change |
|---|---|---|---|---|---|---|
| rs13303327 | ETV5 | G | A | 7.61E-04 | 1.52E-05 | 49.96 |
| rs13303327 | OLIG2 | G | A | 2.11E-02 | 4.53E-04 | 46.65 |
| rs13303327 | TAL1 | G | A | 6.09E-03 | 1.60E-04 | 38.16 |
| rs13303327 | TYY1 | G | A | 1.47E-04 | 5.98E-06 | 24.66 |
| rs13303327 | ELF5 | G | A | 6.56E-04 | 2.66E-05 | 24.64 |
| rs13303327 | ELF2 | G | A | 2.08E-03 | 8.70E-05 | 23.91 |
| rs13303327 | NDF1 | G | A | 4.89E-03 | 2.06E-04 | 23.71 |
| rs13303327 | NFAT5 | G | A | 2.00E-03 | 1.08E-04 | 18.50 |
| rs13303327 | ELF1 | G | A | 2.27E-03 | 1.28E-04 | 17.74 |
| rs13303327 | EHF | G | A | 2.39E-03 | 1.35E-04 | 17.74 |
| rs13303327 | NGN2 | G | A | 7.58E-03 | 4.49E-04 | 16.88 |
| rs13303327 | GABPA | G | A | 2.34E-03 | 1.45E-04 | 16.22 |
| rs13303327 | ETV7 | G | A | 2.08E-03 | 1.36E-04 | 15.31 |
| rs13303327 | ELF3 | G | A | 1.81E-03 | 1.31E-04 | 13.83 |
| rs13303327 | ZN816 | G | A | 3.04E-03 | 2.57E-04 | 11.80 |
| rs13303327 | E2F6 | G | A | 1.47E-03 | 2.05E-04 | 7.17 |
| rs13303327 | ZFP42 | G | A | 5.65E-04 | 8.19E-05 | 6.90 |
| rs13303327 | ETS2 | G | A | 1.70E-03 | 3.02E-04 | 5.65 |
| rs13303327 | ERG | G | A | 3.47E-04 | 6.65E-05 | 5.22 |
| rs13303327 | PLAG1 | G | A | 1.13E-04 | 2.20E-05 | 5.11 |
| rs13303327 | SPI1 | G | A | 1.92E-03 | 3.79E-04 | 5.06 |
| rs13303327 | TYY2 | G | A | 1.19E-05 | 2.43E-06 | 4.89 |
| rs13303327 | FLI1 | G | A | 4.58E-04 | 9.61E-05 | 4.77 |
| rs13303327 | ETS1 | G | A | 4.77E-04 | 1.01E-04 | 4.72 |
| rs13303327 | TAF1 | G | A | 2.36E-05 | 5.21E-06 | 4.54 |
| rs13303327 | TBX15 | G | A | 3.28E-05 | 1.60E-04 | 0.21 |
| rs13303327 | THA11 | G | A | 4.36E-04 | 2.27E-03 | 0.19 |
| rs13303327 | ZN770 | G | A | 3.30E-04 | 1.77E-03 | 0.19 |
| rs13303327 | ZN263 | G | A | 2.33E-06 | 1.28E-05 | 0.18 |
| rs13303327 | ZN143 | G | A | 2.15E-04 | 1.33E-03 | 0.16 |
| rs13303327 | EGR1 | G | A | 1.64E-04 | 1.03E-03 | 0.16 |
| rs13303327 | RREB1 | G | A | 2.93E-04 | 2.16E-03 | 0.14 |
| rs13303327 | Z324A | G | A | 1.15E-04 | 9.57E-04 | 0.12 |
| rs13303327 | ZN140 | G | A | 1.63E-04 | 1.61E-03 | 0.10 |
| rs13303327 | ZN320 | G | A | 7.57E-05 | 7.54E-04 | 0.10 |
| rs13303327 | ZN281 | G | A | 7.57E-05 | 7.61E-04 | 0.10 |
| rs13303327 | ZNF76 | G | A | 5.79E-05 | 5.82E-04 | 0.10 |
| rs13303327 | WT1 | G | A | 7.60E-06 | 8.20E-05 | 0.09 |
| rs13303327 | KLF15 | G | A | 4.99E-08 | 5.49E-07 | 0.09 |
| rs13303327 | EGR2 | G | A | 5.84E-05 | 7.54E-04 | 0.08 |
| rs13303327 | ZN263 | G | A | 9.79E-06 | 1.47E-04 | 0.07 |
| rs13303327 | PURA | G | A | 1.06E-06 | 6.52E-05 | 0.02 |

| | | | | | | |
|---|---|---|---|---|---|---|
| rs13303160 | FOSL1 | G | A | 4.32E-04 | 4.94E-06 | 87.31 |
| rs13303160 | JUND | G | A | 9.47E-04 | 2.28E-05 | 41.63 |
| rs13303160 | BATF | G | A | 1.08E-02 | 2.65E-04 | 40.95 |
| rs13303160 | FOSB | G | A | 1.18E-03 | 2.93E-05 | 40.11 |
| rs13303160 | FOSL2 | G | A | 8.16E-04 | 2.15E-05 | 37.93 |
| rs13303160 | JUNB | G | A | 8.00E-04 | 2.47E-05 | 32.36 |
| rs13303160 | JUN | G | A | 8.40E-04 | 2.68E-05 | 31.37 |
| rs13303160 | ZFX | G | A | 1.90E-03 | 7.88E-05 | 24.15 |
| rs13303160 | FOS | G | A | 5.73E-04 | 2.74E-05 | 20.86 |
| rs13303160 | BACH2 | G | A | 8.84E-04 | 8.59E-05 | 10.29 |
| rs13303160 | MAFK | G | A | 2.19E-03 | 2.33E-04 | 9.38 |
| rs13303160 | BACH1 | G | A | 1.80E-04 | 2.85E-05 | 6.30 |
| rs13303160 | ZN554 | G | A | 6.28E-04 | 1.11E-04 | 5.65 |
| rs13303160 | NFE2 | G | A | 3.72E-04 | 7.80E-05 | 4.77 |
| rs13303160 | ZN329 | G | A | 1.08E-03 | 2.35E-04 | 4.58 |
| rs13303160 | PAX2 | G | A | 1.68E-04 | 4.00E-05 | 4.19 |
| rs13303160 | MBD2 | G | A | 4.22E-06 | 2.85E-05 | 0.15 |
| rs13303160 | ATF6A | G | A | 3.91E-04 | 4.34E-03 | 0.09 |
| rs13303160 | THAP1 | G | A | 1.09E-04 | 1.34E-03 | 0.08 |
| rs13303160 | HEY2 | G | A | 1.58E-04 | 1.96E-03 | 0.08 |
| rs13303160 | HES7 | G | A | 1.08E-04 | 1.53E-03 | 0.07 |
| rs13303160 | HES5 | G | A | 4.58E-04 | 7.07E-03 | 0.06 |
| rs13303160 | HEY1 | G | A | 2.65E-04 | 7.97E-03 | 0.03 |

Predicted TF motifs disrupted by SNP alleles for either rs13303327 or rs13303160. Alleles 1 and 2 are defined and *P*-values of predicted binding strength are indicated for each allele. The fold-change between the two *P*-values was calculated and used to determine the best predictions.
