## Supplementary Figure 6 for "Allelic effects on *KLHL17* expression likely mediated by JunB/D underlie a PDAC GWAS signal at chr1p36.33"

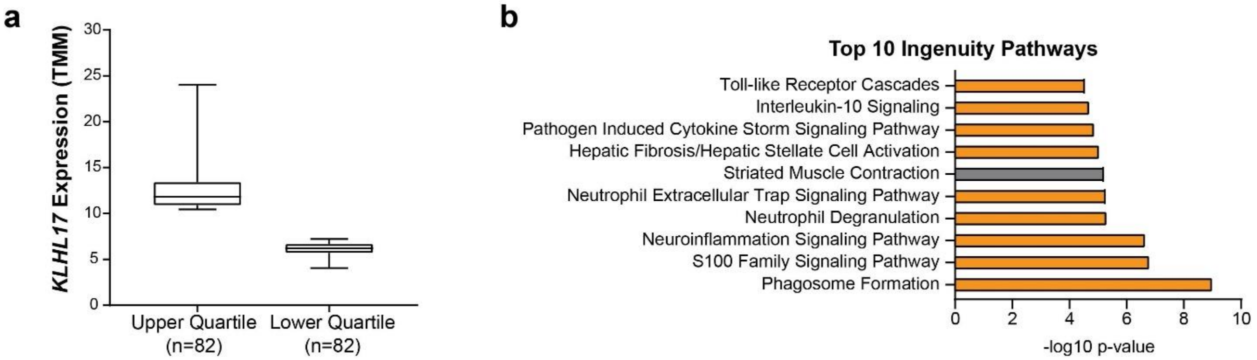


**Supplementary Figure 6: *In silico KLHL17* knockdown quartiles and Ingenuity Pathway Analysis (IPA)** a) Boxplot indicating the *KLHL17* expression (TMM) of GTEx Pancreas samples in the upper (75%) and lower (25%) quartiles of samples; b) The top ten most significant pathways from the in silico differential gene expression analysis identified in Ingenuity Pathway Analysis plotted based on the -log10 *P* value. Orange bars indicate a pathway associated with inflammation.
