## Supplementary Figure 5 for "Allelic effects on *KLHL17* expression likely mediated by JunB/D underlie a PDAC GWAS signal at chr1p36.33"

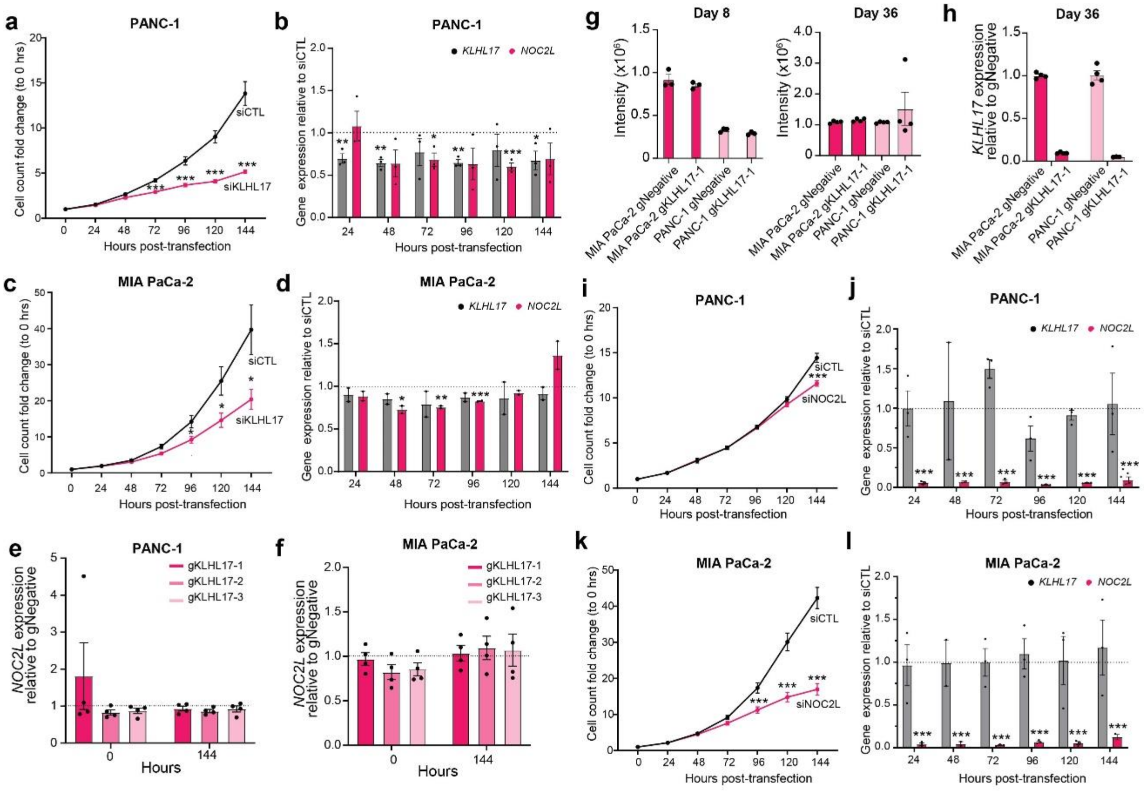


**Supplementary Figure 5: *In vitro* growth analysis using siRNA mediated knockdown for *KLHL17* and *NOC2L*** a,c) Cell count normalized to 0 hours post-transfection of non-targeting (siCTL) or *KLHL17* siRNA in the PANC-1 (n=10) and MIA PaCa-2 (n=8) cell lines, respectively; b, d) qPCR analysis of the knockdown efficiency for both *KLHL17* (grey) and nearby *NOC2L* (pink) relative to siCTL and internal *HPRT* control in PANC-1 (n=3) and MIA PaCa-2 (n=2) cells over the course of the growth assay, respectively. ; e, f) qPCR analysis of *NOC2L* expression in the CRISPRi-mediated *KLHL17* knockdown at the beginning and end of the growth analysis in Figure 5 for PANC-1 and MIA PaCa-2 cell lines, respectively. Expression is relative to the sgNegative control and internal control *HPRT*; g) Peptide intensities for global proteomic analysis at days 8 and 36 in MIA PaCa-2 and PANC-1 gNegative and gKLHL17 cells. n=3 for Day 8 and n=4 for Day 36; h) Corresponding qPCR for KLHL17 at Day 36 normalized to the gNegative and internal *HPRT* control. n=1 biological replicate. Dots represent technical replicates. i,k) Cell count normalized to 0 hours post-transfection of non-targeting (siCTL) or NOC2L siRNA in the PANC-1 (n=12) and MIA PaCa-2 (n=12) cell lines, respectively; j, l) qPCR analysis of the knockdown efficiency for both *KLHL17* (grey) and nearby *NOC2L* (pink) relative to siCTL and internal *HPRT* control in PANC-1 and MIA PaCa-2 cells over the course of the growth assay, respectively; k) For all graphs, error bars represent the SEM. Unpaired, two-tailed t-tests were performed. * $P < 0.05$, ** $P < 0.01$ *** $P < 0.001$
