## Supplementary Figure 4 for "Allelic effects on *KLHL17* expression likely mediated by JunB/D underlie a PDAC GWAS signal at chr1p36.33"

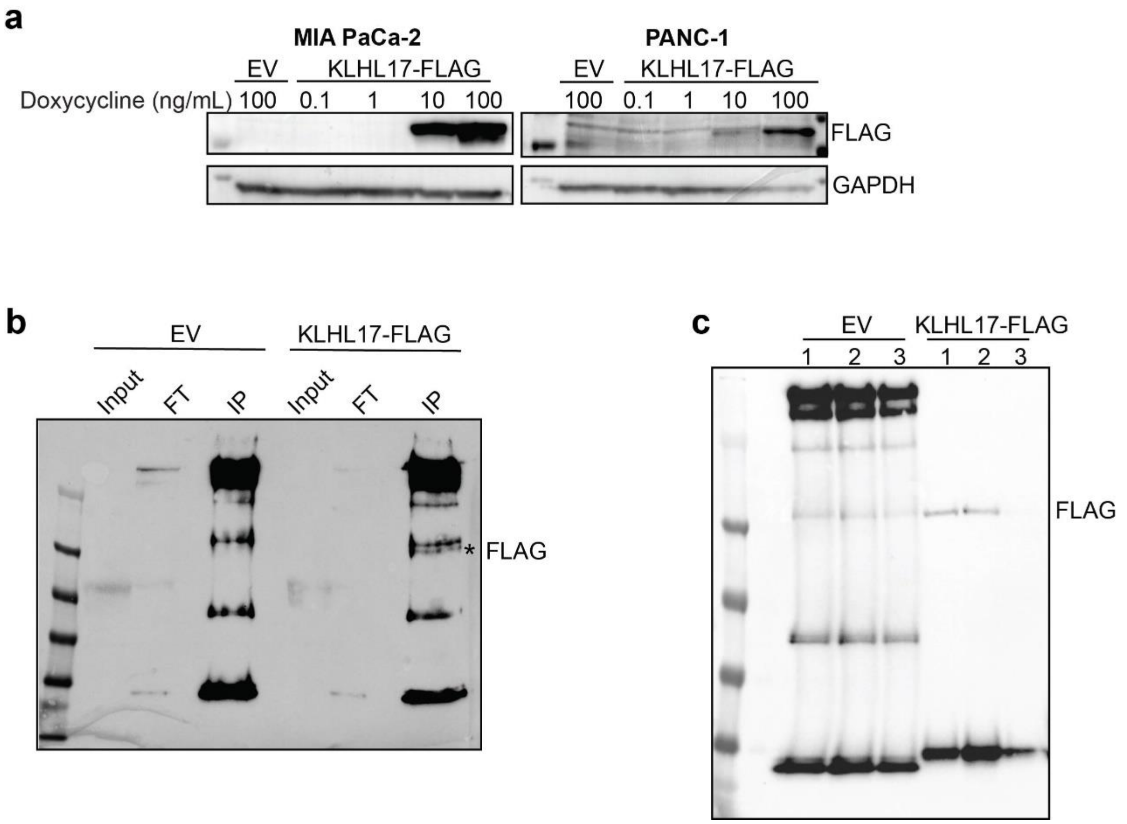


**Supplementary Figure 4: Western blot analysis of *KLHL17* overexpression and immunoprecipitation** a) Western blot using whole cell lysate from MIA PaCa-2 and PANC-1 cells that were stimulated with increasing amounts of doxycycline for 72 hours to induce KLHL17-FLAG expression. Empty Vector (EV) was used as a negative control for FLAG expression. FLAG antibody was used for detection of induced KLHL17 and GAPDH antibody was used for loading control; b) Western blot of the FLAG immunoprecipitation from EV or KLHL17-FLAG expressing PANC-1 cells that was used for a pilot mass-spectrometry run to identify co-immunoprecipitated proteins. Blot was probed with FLAG antibody; c) Western blot of 3 FLAG immunoprecipitation replicates from EV or KLHL17-FLAG expressing PANC-1 cells for mass spectrometry. Blot was probed with FLAG.
