## Supplementary Figure 3 for "Allelic effects on *KLHL17* expression likely mediated by JunB/D underlie a PDAC GWAS signal at chr1p36.33"

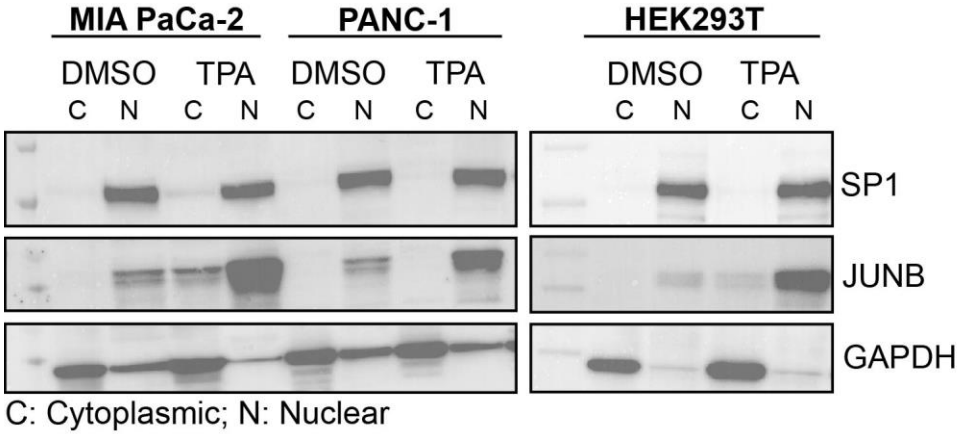


**Supplementary Figure 3: Western blot confirmation for induction of AP1 protein expression with TPA treatment.** Western blot analysis using cytoplasmic and nuclear extracts from MIA PaCa-2, PANC-1 and HEK293T cells following 48-hour treatment of cells with DMSO or TPA. Antibodies for SP1 and GAPDH were used as loading controls for nuclear and cytoplasmic extracts, respectively.
