## Supplementary Figure 2 for "Allelic effects on *KLHL17* expression likely mediated by JunB/D underlie a PDAC GWAS signal at chr1p36.33"

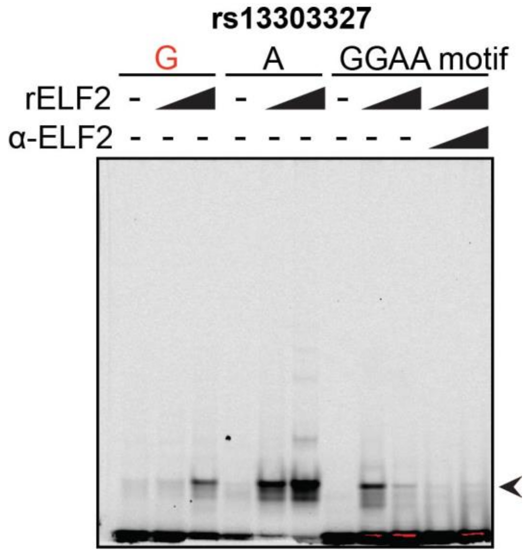


**Supplementary Figure 2: Validation of ELF2 binding *in vitro*.** EMSA using recombinant ELF2 and the rs13303327 sequence or a control sequence with the ELF GGAA motif centered amongst scrambled nucleotides. Increasing amounts of ELF2 recombinant protein were used for each oligo and with the control sequence increasing amounts of ELF2 antibody was included. The risk allele is indicated in red. The arrow denotes the allele-specific binding bind.
