## Supplementary Figure 1 for "Allelic effects on *KLHL17* expression likely mediated by JunB/D underlie a PDAC GWAS signal at chr1p36.33"

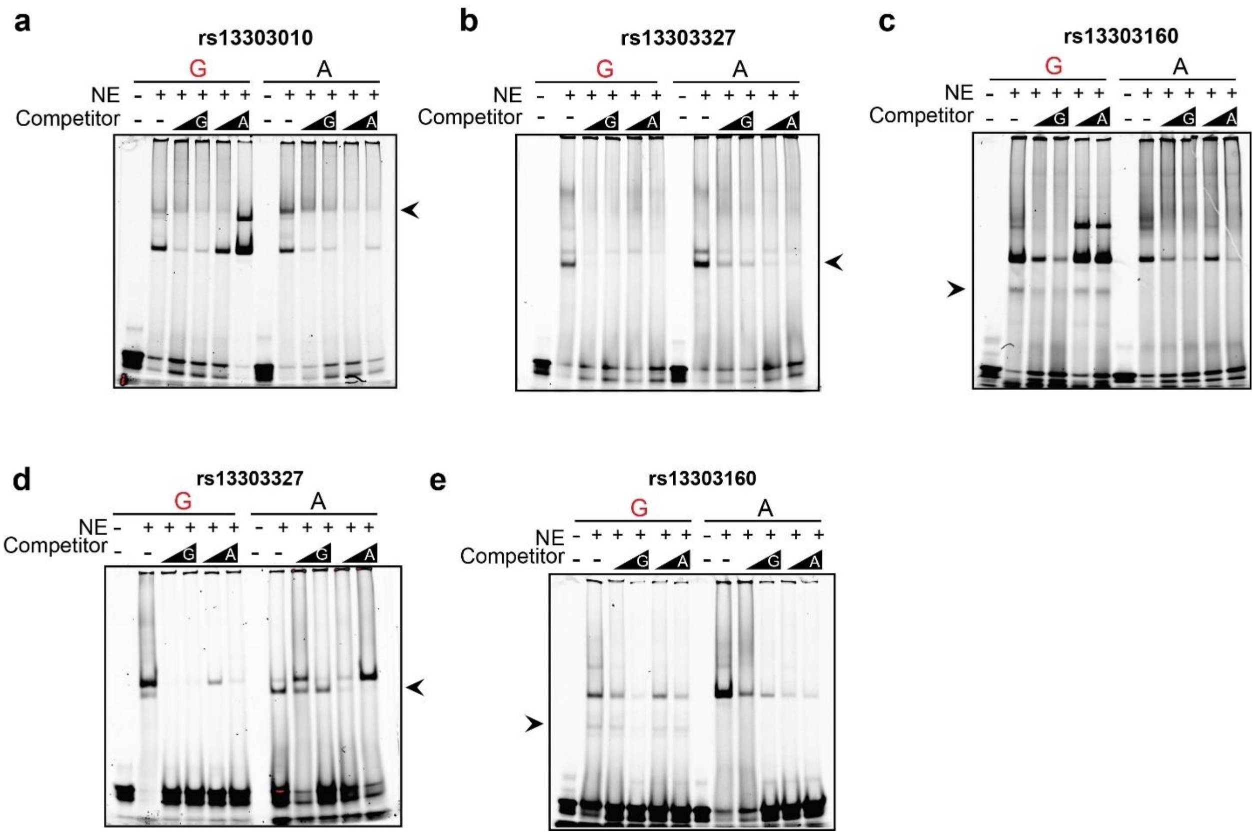


**Supplementary Figure 1: Confirmation of allele-specific binding *in vitro*** a-c) Representative EMSA with MIA PaCa-2 nuclear extract and fluorescently labeled oligonucleotides for rs13303010, rs13303327, rs13303160, respectively. Competitor is the same sequence with no fluorescent label in excess (50, 100X); d, e) EMSA with HeLa extract for rs13303327 and rs13303160, respectively. Risk alleles are indicated in red.
